# Adaptive data-driven selection of sequences of biological and cognitive markers in pre-clinical diagnosis of dementia

**DOI:** 10.1101/2021.10.26.21265515

**Authors:** Patric Wyss, David Ginsbourger, Haochang Shou, Christos Davatzikos, Stefan Klöppel, Ahmed Abdulkadir, the ISTAGING Study, the Alzheimer’s Disease Neuroimaging Initiative, the Australian Imaging Biomarkers and Lifestyle flagship study of ageing

## Abstract

Effective clinical decision procedures must balance multiple competing objectives such as time-to-decision, acquisition costs, and accuracy. We describe and evaluate POSEIDON, a data-driven method for PrOspective SEquentIal DiagnOsis with Neutral zones to individualize clinical classifications. We evaluated the framework with an application in which the algorithm sequentially proposes to include cognitive, imaging, or molecular markers if a sufficiently more accurate prognosis of clinical decline to manifest Alzheimer’s disease is expected. The algorithm chose to include optional invasive markers in 37 percent of cases at the cost of 1 percent lower accuracy. Applied to longitudinal data, POSEIDON selected 14 percent of all available measurements and concluded after an average follow-up time of 0.74 years at the expense of five percent lower accuracy. While effective in obtaining timely and economical decisions, our multi-objective evaluation implies that the implementation into consequential clinical applications remains controversial because of the intrinsic dependence on inherently subjective prescribed cost parameters.

## Introduction

Timely and correct diagnosis of dementia due to Alzheimer’s disease (AD) improves treatment and reduces care costs.^1^ Diagnostic uncertainty—even in specialized centers, however, is high. This results in sensitivity ranging from 71 to 87 percent and specificity ranging from 44 to 71 percent^2^ but follow-up examinations and invasive exams improve accuracy. Thus, to date, a typical diagnostic decision of dementia is based on a panel of cross-sectional or a sequence of repeatedly measured (longitudinal) markers from multiple modalities such as magnetic resonance imaging (MRI) or cognitive testing.^3–5^ There is currently no consensus or systematic approach to individualize the selection of panels and temporal sequences of markers to acquire. Herein, we present a data-driven framework for PrOspective SEquentIal DiagnOsis with Neutral zones (POSEIDON) that integrates irregularly sampled, repeated (longitudinal), multi-variate data with varying numbers of observations and derives an individually adaptive expansion of the panel of markers for classification as exemplified on **Fig. 1**. While the core method is generic, it is evaluated in this study with an implementation of a parametric multi-variate linear mixed-model based classifier to predict progression from mild cognitive impairment to manifest Alzheimer’s disease.

**Fig. 1:**
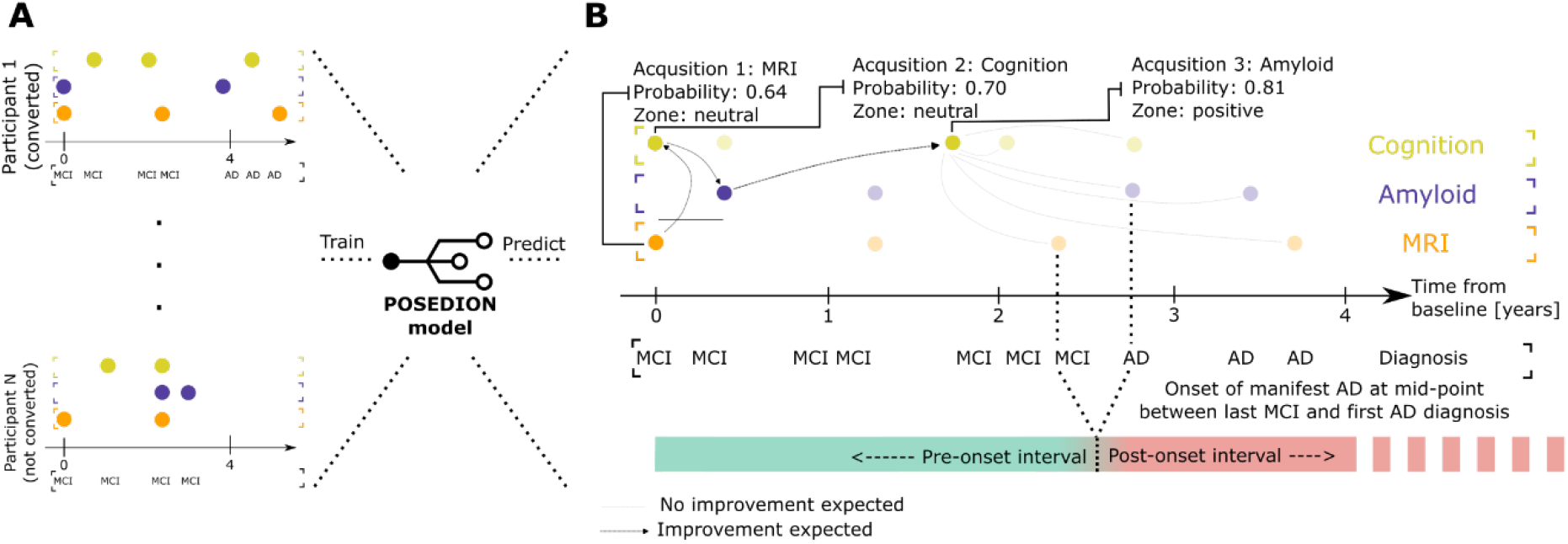
Example application of POSEIDON. Example of the application of POSEIDON to a set of retrospectively acquired markers of cognition, brain MRI, and Amyloid of a single participant. The task was to predict the conversion from MCI to AD within three years from baseline. The mid-point between the last diagnosis MCI and the first diagnosis of AD was defined as time of conversion; 2.6 years in this example. Solid colored disks indicate measurements taken and used for training or prediction, whereas opaque colored disks indicate available measurements that were not observed by the sequential algorithm. At each stage, the algorithm opts to observe one of the proposed measurements at the same or later time, or concludes the decision process with a definitive prediction. In the shown example, MRI was selected first (based on age), followed by cognition at the same visit. The next variable selected was Amyloid at the four-month visit. Then, the algorithm skipped potential exams at the 1.3-year mark and concluded the prediction with a second cognitive test 0.4 years later as none of the examinations afterwards was expected to increase the accuracy more than the cost they would incur. Of note, while all retrospectively acquired measurement were known within the context of the evaluation procedure, the algorithm itself was only given the information of variable type and time during the selection process.

Unlike in settings in which the diagnosis is based on fixed sets of measurements,^6^ we mimic a clinically more relevant setting in which the sequence of markers—that is which marker is acquired when, is not set a priori but instead individualized based on data-driven modelling. To implement the framework, the task was formulated as a sequential classification task with a neutral zone. Neutral zone classifiers^7–9^ have a decision rule have a neutral outcome in addition to the positive and negative label of a forced choice classifier. To perform a sequential classification task, a selection rule is required to choose which measurement to include next. Individualizing the panel of markers with a decision and selection rule requires to balance multiple competing targets such as accuracy, patient burden, financial costs and time to diagnosis. Loosely worded, the multi-faceted objective is to reach an early, accurate diagnosis with little resources and limited patient burden. The relative importance of these aspects are tuned and compared across strategies by prescribed cost parameters that are set *a priori*.

For our evaluation, we focus on a diverse data set of four markers as predictors of clinical progression in AD that capture pathological hallmarks, AD-like brain atrophy, and cognitive markers. The invasive A*β*_1−42_ cerebro-spinal fluid (CSF) marker^10^ imposes a high burden on patients and high monetary costs. These shortcomings are compensated by higher sensitivity of the prognosis. Conversely, the two chosen cognitive assessments Mini-Mental-State Examination (MMSE)^11^ and Rey Auditory Verbal Learning (RAVLT)^12^ have a lower economic cost and patient burden, but also a lower accuracy in early stages of the disease. Non-invasive magnetic resonance imaging (MRI) provides machine-learning derived measures of AD-like atrophy (SPARE-AD)^13^ that have intermediate cost of acquisition, intermediate sensitivity, and high specificity for typical amnestic AD.

Our implementation of a sequential neutral zone classifier that leverages models of potential prospective gains in accuracy is based on an empirical Bayes approach that embeds multi-variate linear mixed-effects models in the context of classification and accounts for dependencies between markers and their trajectories. The mixed-effects classification approach is more flexible in handling irregularly sampled data and models longitudinal data with fewer parameters than other generative approaches such as traditional discriminant analysis.^10–14^ Moreover, estimating underlying distributions of markers—rather than only their effect on decision values (such as e.g., the posterior probability), allows to quantify the impact of future measurements, conditional on the already completed assessments (see Supplementary **Fig. S1** for a visualization of different types of classifiers). For each individual, POSEIDON sequentially proposes to include optional markers only if the total decision costs are expected to decrease. POSEIDON is useful to calibrate the trade-off between competing goals of the prognostic process selecting a limited panel of markers that empirically lowers the quantitative objective with respect to a diagnosis based on the already complete panel of markers. The individualization of the process is expected to manifest in a better (depending on the *a priori* prescribed costs) balance of accuracy, time to diagnosis, and used resources than classification based on fixed panels of measurements. Additional markers would only be acquired in those uncertain cases where an increase in accuracy outweighs measurement costs given by the acquisition and the delay of the decision. Thus, if and which markers to include depends on past acquisition as well as options for future acquisitions. In the limit cases of exceedingly high and low prescribed costs for additional measurements, we expect an accuracy equivalent to no or all additional measurements, respectively. A consequence of the multi-faceted evaluation of performance is that there is no straight-forward unequivocal measure of superiority across diagnostic procedures. One procedure may outperform another procedure in some aspects (for example accuracy), but perform worse in others (for example number of acquisitions). Nevertheless, some strategies based on fixed panels may be dominated by others in all aspects. We sought to identify sequential algorithms that are competitive in all aspects and characterize the effect of costs. Given the heterogeneity of disease effects on brain morphometry, amyloid burden, and cognitive outcome, we investigated the effect of this heterogeneity on misclassification rates. As a proxy for clinical usefulness for disease prognosis, we evaluated the pre-manifest sensitivity as an additional objective metric which we defined as the ratio of participants that were correctly identified as converters before the onset of symptoms.

## Results

### Selective inclusion of invasive measurements increases sensitivity

Here, we evaluated the performance in a scenario in which all participants would receive an MRI and then based on the outcome of the classification with SPARE-AD would either be definitively classified or referred to a lumbar puncture procedure to obtain A*β*_1-42_ - CSF. The sequential classifier that optionally included the A*β*_1-42_ - CSF measurement conditional on the observed baseline MRI and age had lower total cost when low measurement costs were prescribed, equal total costs when high measurement costs were prescribed, and slightly higher total costs in three cases where intermediate costs were prescribed (**Fig. 2A**).. In case only MRI was used for all participants, the mean total cost was equal to the 27 percent error rate, while for classifications always using both biomarkers the mean total costs correspond to the error percentage of 20 plus the prescribed measurement costs for A*β*_1-42_ - CSF. Thus, the intersection between the lines of always MRI and always MRI+ A*β*_1-42_ - CSF was where the measurement cost of A*β*_1-42_ - CSF was set to 7. Lowering prescribed costs for measuring A*β*_1-42_ - CSF coincided with an increase in accuracy and an increase in the fraction of acquisition of A*β*_1-42_ CSF (**Fig. 2B**). The increase in accuracy was mainly driven by an increase in sensitivity (structural MRI alone: accuracy of 0.73, specificity of 0.78 and sensitivity of 0.65, inclusion of A*β*1-42-CSF for all cases: accuracy of 0.80, specificity of 0.80 and sensitivity of 0.80) without reduction in specificity (**Fig. 2C-D**). Accuracies of the sequential classifiers approached the one using both measures in all cases even when including A*β*_1-42_ - CSF in less than 50 percent of the cases. For example, 98 percent of maximum accuracy (98 percent of specificity and 98 percent of sensitivity) was achieved with 37 percent of A*β*_1-42_ - CSF measures. The misclassification cost parameter was fixed as 100 for all analyses (see Supplementary Methods 1 for detailed information about decision costs) leading to measurement costs of A*β*_1-42_ CSF that are given as percentage of the costs of one misclassification. For measurement costs of x the A*β*_1-42_ CSF is included if the expected increase in accuracy is higher than x/100 (see Supplementary Equation (S13)). The considered data consisted of 410 subject (167 MCI-converters, see Supplementary **Fig. S2**).

**Fig. 2:**
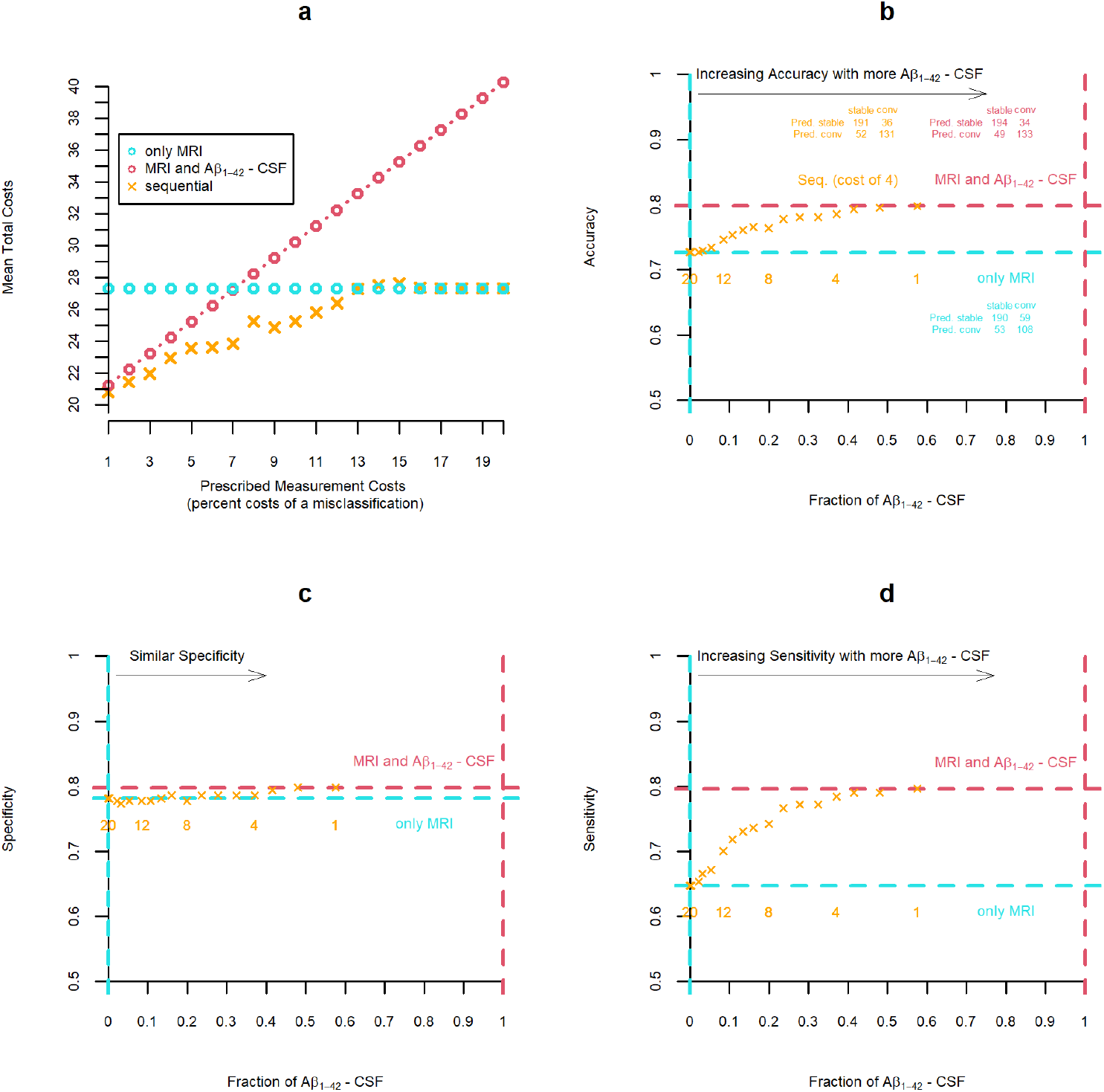
Two-stage classification. Multi-facetted comparison of sequential two-stage classifier and classifications based on fixed panels of measurements (only MRI or always MRI and A*β*_1-42_ - CSF for all subjects) classifiers for varying measurement costs (1 to 20). Note that the scale of the y-axes in a, b, and c start at 0.5 (chance level) and not at 0 (minimum possible value). **(a)**: Mean total cost resulting from varying prescribed measurement cost of A*β*_1-42_ - CSF obtained with sequential and non-sequential strategies. **(b)** – **(d)**: Portion of all cases for which A*β*_1-42_ - CSF was included and resulting accuracy **(b)**, specificity **(c)** or sensitivity **(d)**.

### Identifying cases for which prognosis with structural MRI alone is uncertain

We used SPARE-AD derived from MRI and a fixed prescription of measurement costs (c=4) of A*β*_1-42-_ CSF to split the sample into confident prognoses (definitively predict either MCI-stable or MCI-converter with a sequential classifier, 258 participants, 86 of which were MCI-converters) and uncertain prognoses (predict “neutral zone” with two-stage classifier, 152 participants, 81 of which were MCI-converters). When including all participants, the accuracy for a classification with MRI was 0.73. Based on SPARE-AD, 55 percent of all uncertain prognoses and 83 percent of all confident prognoses were correct. When updating the uncertain predictions by adding A*β*_1-42_ -CSF the percentage of correct classification increased to 71(+16 percent).

As illustrated in **Fig. 3**, including A*β*_1-42-_ CSF additional to the MRI measurements led to higher sensitivity (+0.15) with similar specificity (+0.02), while including all longitudinal cognitive measurements (that covers also measurements after the conversion to manifest AD) additional to the two cross-sectional biomarkers was beneficial for both specificity and (accuracy: +0.08, specificity: +0.06, sensitivity: +0.11). Classification based on MRI only produced a similar number of false positive cases as classification with both MRI and A*β*_1-42-_ CSF (22 respectively 20 percent of MCI-stables). The overlap was 15 percent of all MCI-stables (**Fig. 3A**). From the 49 false positive predictions with MRI and A*β*_1-42-_ CSF 24 could correctly be classified as MCI-stables when additionally, the progression of the cognitive markers was considered for prediction. For the leftover 25 false positive cases the raw data was examined to identify why they are MCI-stables (see Supplementary **Fig. S3**). Moreover, as displayed in **Fig. 3B** the inclusion of A*β*_1-42_ -CSF for classification led to 23 fewer uncertain cases that were falsely classified as negative (0.14 reduction of false negative rate) but only 2 fewer easy cases (0.01 reduction of false negative rate).

**Fig. 3:**
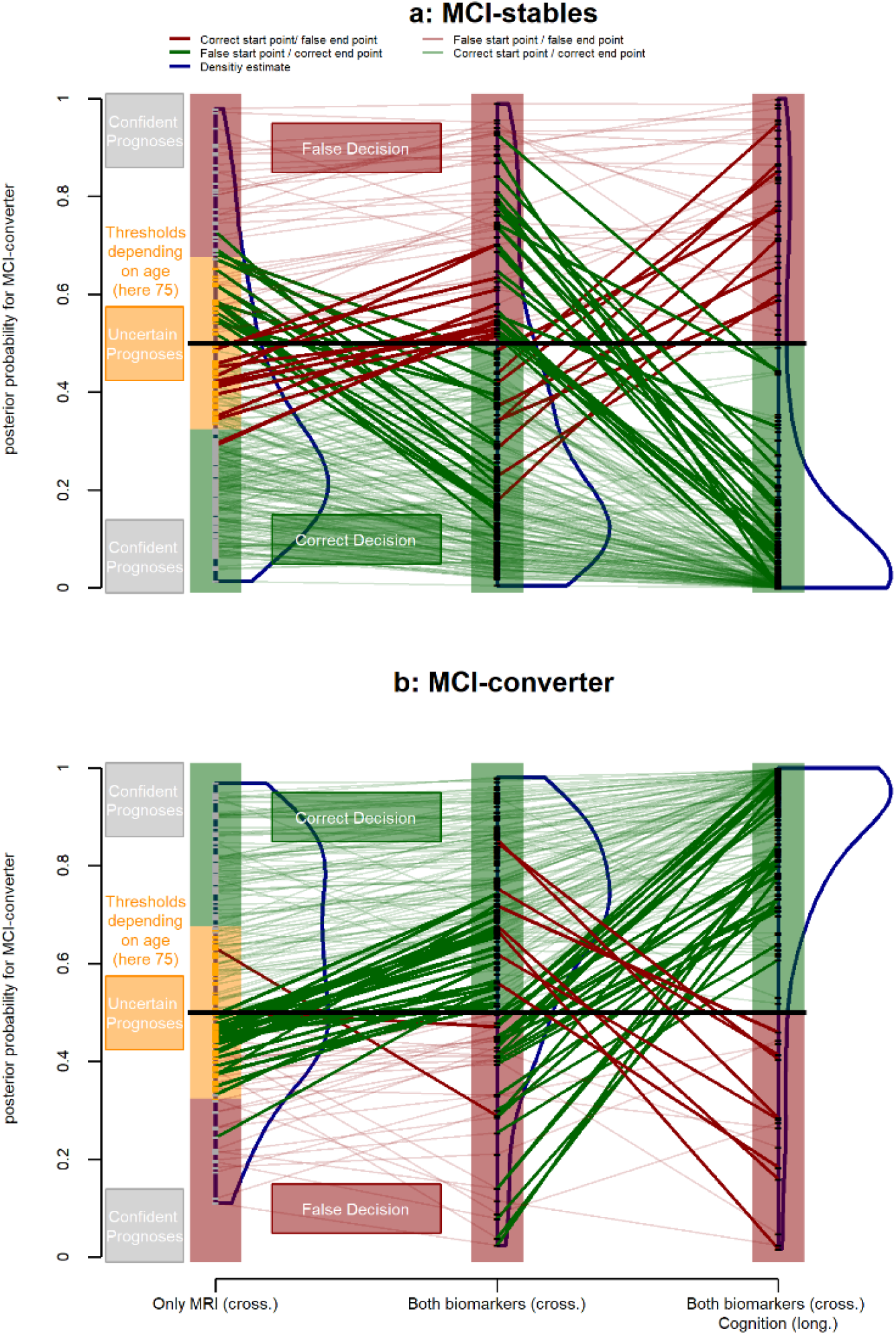
Chart displaying posterior probabilities belonging to the population of MCI-converters of 410 participants either predicted with cross-sectional MRI measure only, cross-sectional MRI and A*β*_1-42-_ CSF measures or the cross-sectional MRI and A*β*_1-42_ -CSF as well as all cognitive measures (MMSE and RAVLT measures of all time points) separately for all MCI-stables **(a)** or MCI-converters **(b)**. Regions of predictions of (1) a sequential classifier based on cross-sectional MRI (optional A*β*_1-42_ -CSF for uncertain subjects with neutral prediction) in the left column, (2) a classifier with both MRI and A*β*_1-42_ -CSF in the middle column or (3) a classifier with the fixed panel including both cross-sectional biomarkers and longitudinal cognitive markers in right column. For classification with MRI also neutral predictions (initially uncertain cases) are possible. Connected dots indicate that these belong to the same participant. Thick lines indicate that the classification changed, thin lines indicate no change. Green lines indicate correct and red lines incorrect classification with the posterior probability of the lines endpoint.

In **Fig. 4**, analyses covering the Amyloid (A)-Tau (T)-Neurodegeneration (N) status ^14^ is shown for 301 participants (99 of them MCI-converters) for which the complete A/T/[N] classification was available The distribution of A/T/N profiles were similar for MCI-stables and negatively predicted cases as well as MCI-converters and positively predicted cases (**Fig. 4c-d**). On one hand, the sequential classification and the classification based on MRI and A*β*_1-42-_ CSF showed less false negatives for profiles with A+ and T+ compared to a classifier based on MRI only, but the same number for all other profiles (**Fig. 4e**). For one patient with A+/T+/N- and one patient with A+T+N+, the sequential classifier missed to include the A*β*_1-42-_ CSF that would have led to a correct classification. On the other hand, including A*β*_1-42-_ CSF additional to MRI led to higher number of false positive classifications of patients with an A+/T+ profile and a lower number of false positive predictions of patients with A-profile (**Fig. 4f**).

**Fig. 4:**
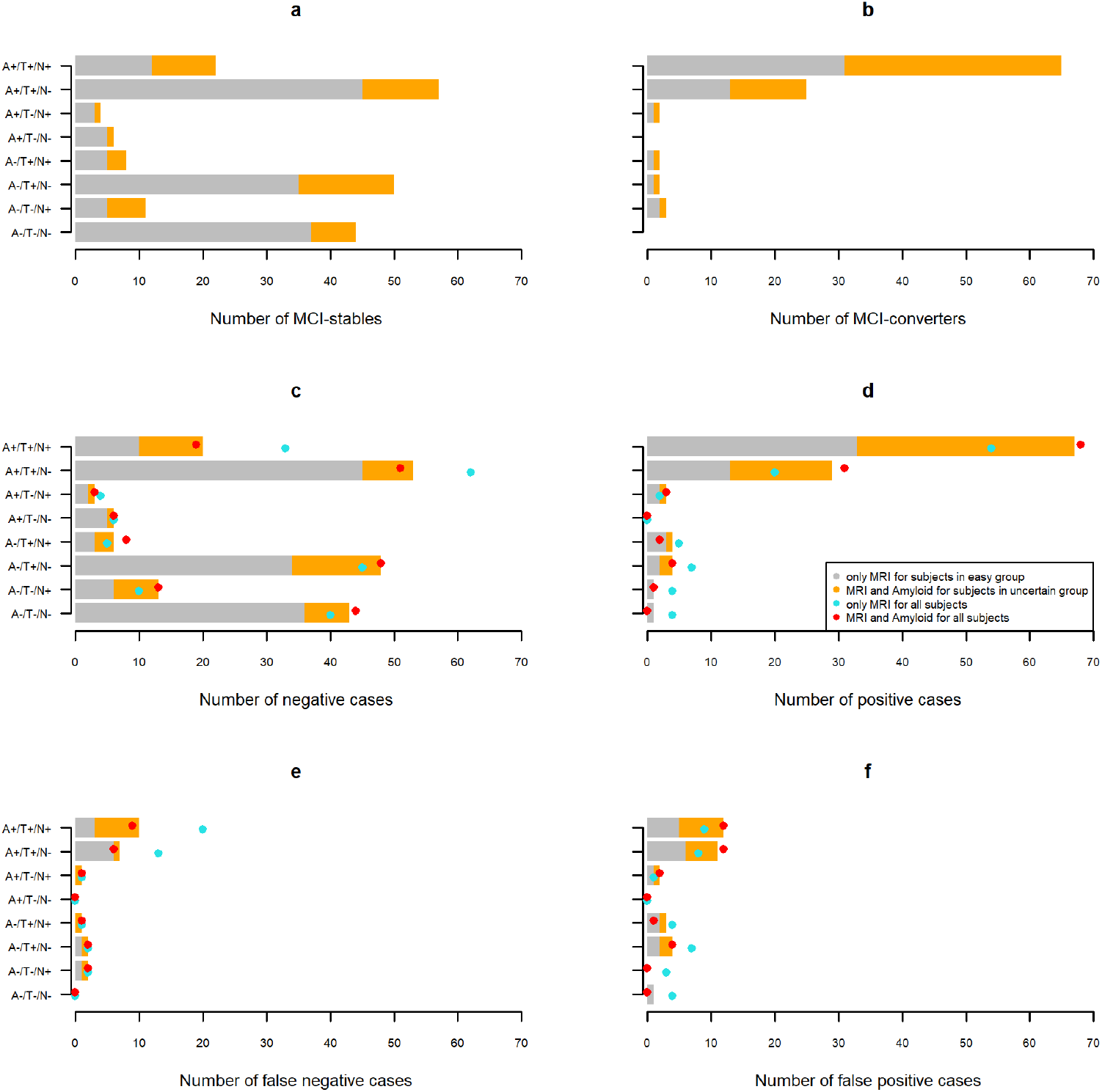
Distribution of A/T/N profiles of easy (grey) and uncertain (orange) cases. **(a)** - **(b)**: Distributions for MCI-stables respectively MCI-converters. **(c)** - **(d)**: Distributions of positive/negative classifications. For the sequential classifier the number of cases are separated into the number of easy (grey bar, classified with MRI) and uncertain (orange bar, classified with MRI and A*β*_1-42_ - CSF) cases. The blue dots represent the positively/negatively predicted cases when using MRI only for the whole sample and the red dot the positively/negatively predicted cases using MRI and A*β*_1-42_ - CSF for the whole sample. **(e)** - **(f)**: As in **(c)** – **(d)** but displaying only the number of false positive/negative cases instead of all positive/negative cases.

Predictions based on MRI only led to more distinct survival curves respectively higher differences in the hazard rates when fitted on easy cases than when fitted on uncertain **Fig. 5**. When the A*β*_1-42-_ CSF measure was included in uncertain cases, the survival curves and hazard rates of the ones predicted as MCI-converter and the ones predicted as MCI-stables became more similar to the ones predicted for easy cases based on MRI only. Details about estimation techniques, additional results, and exploratory testing for significant differences between groups are reported in the Supplementary Methods 5 and Supplementary Results 1.

**Fig. 5:**
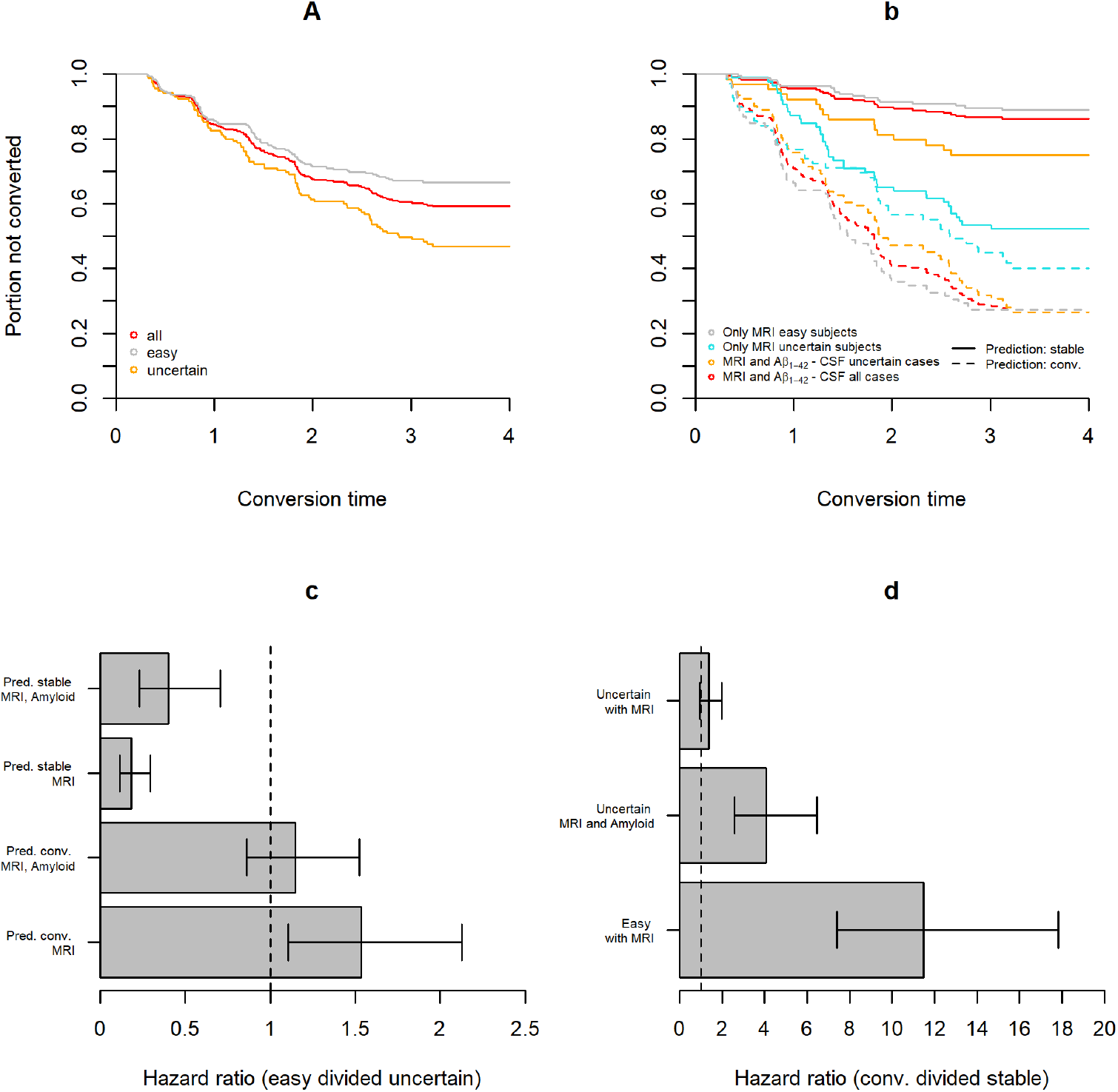
Survival analysis and hazard ratios. Survival curves showing the portion of not progressed participants estimated with the Kaplan Meier technique and hazard ratios estimated with Cox-regressions. **a)**Survival curves fitted only on easy cases, only uncertain cases, or the whole sample. **b** Survival curves fitted separately on participants predicted not to progress to AD or participants predicted to convert to AD by different classifiers and split by easy/uncertain. Classifiers either used only the MRI measure or both the MRI and A*β*_1-42_ -CSF measure for prediction. **c** Ratio of the hazard rate of easy cases divided by the hazard rate of uncertain cases separately for participants predicted not to progress to AD or to convert to AD. For easy cases only the MRI is used for classification while for uncertain cases either only the MRI or the MRI and A*β*_1-42_ -CSF measures are considered for classification. Error bars correspond to lower and upper bounds of a 95% confidence interval (estimated via Wald’s method, see Supplementary Equation S25). **d** Ratio of the hazard rate of cases predicted to convert to AD divided by the hazard rate of cases predicted not to progress to AD by different classifiers and split by easy/uncertain (error bars are 95% confidence intervals as in **c**).

### Balancing accuracy, number of assessments, and time to diagnosis

Sequential classifiers that balance accuracy, number and type of measurements, and the time to decision showed lower mean total costs than fixed strategies such as uni- and multi- variate cross- sectional - and longitudinal strategies for a wide range of cost parameters (see Supplementary Tab. S1). The sequential classifier either selected the earliest observation with expected costs reduction (greedy strategy) or the observation with the highest expected cost reduction (exhaustive strategy) as next observation to the panel of measurements. As shown in **Fig. 6**a-b, the use of more resources (measurements or time) increased accuracy. Lower prescribed costs of time or acquisitions coincided with lower average time to diagnosis or fewer observations, respectively. Sequential classifiers tuned to favor delaying the diagnosis and/or taking more measurements tended to be more specific and more sensitive (**Fig. 6c-f**). The sequential classifiers approached the maximum accuracy that was achieved by combining all available data. By combining all available data from 20.9 measurements per subject on average that were acquired over 4.8 years on average an accuracy of 0.89, specificity of 0.88, and sensitivity of 0.90 was achieved.

**Fig. 6:**
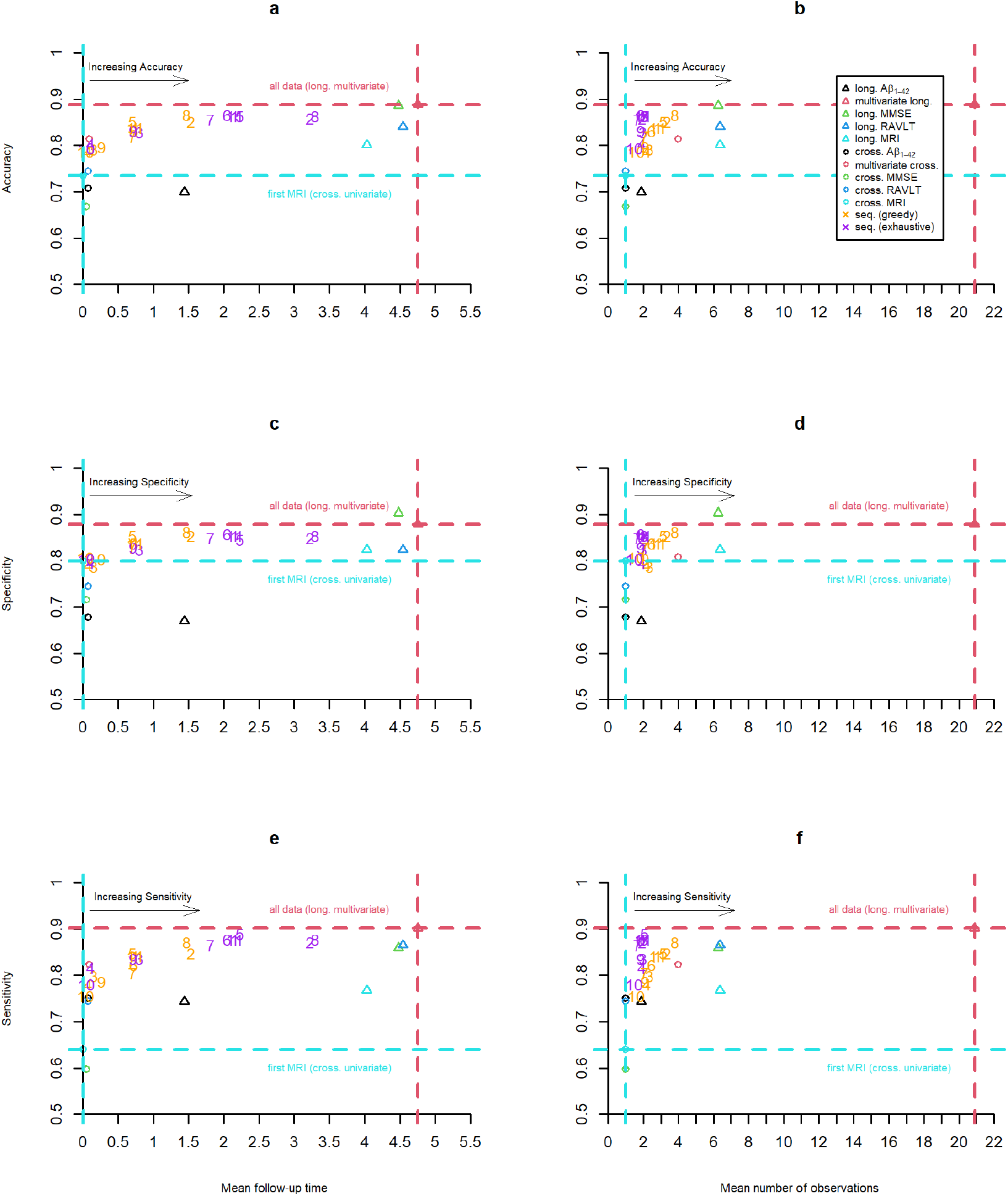
Sequential and non-sequential variable selection strategies. Quantitative comparison of sequential (for varying costs of acquisition and time, see below) and fixed i.e., univariate (one marker) or multivariate (all markers) cross-sectional (cross., only measurement at baseline) or longitudinal (long., all repeated measurement of the markers) strategies. Note that the scale of the y-axes at 0.5 (chance level) and not at 0 (minimum possible value). Mean follow-up time or mean number of observations and resulting accuracy (in **a** and **b**), specificity (in **c** and **d**) or sensitivity (in **e** and **f**) are displayed. Scattered numbers 1 to 10 in the plot correspond to results obtained with tuples of prescribed costs (time; MRI; A*β*1-42 -CSF; cognitive test); 1: (2; 2; 4; 1), 2: (1; 2; 4; 1), 3: (4; 2; 4; 1), 4: (8; 2; 4; 1), 5: (2; 1; 2; 0.5), 6: (2; 4; 8; 2), 7: (2; 8; 16; 4), 8: (1; 1; 2; 0.5), 9: (4; 4; 8; 2)., 10: (8; 8; 16; 4)

For the results presented in this section and the Supplementary Results 2 (of overall 403 participants, 164 MCI-converters) we set misclassification costs to 100 and considered varying marker specific costs of acquisition (includes patients burden and financial costs) and costs per year of waiting (measurement costs of optional observations sum of costs of waiting /delay of decision and costs of acquisition, see Supplementary Methods Section 1). Detailed results covering a wider range of objective metrics such as costs from different sources, performance measures, number of measurements per type, and the time at which they occurred evaluated with varying cost parameters are reported in Supplementary **Tab. S1** (the table includes results covering the quadratic discriminant model as descripted in the Supplementary Methods 3 and 4).

### Multiple objectives: Competitiveness and dominance of decision strategies

Inclusion of more data points or a longer observation interval tended to result in higher classification performance. In a setting with varying number of observations across competing methods, no single metric is sufficient to claim superiority. Multiple objective metrics are needed to evaluate decision strategies in sufficient depth. As shown in **Fig. 6** for accuracy, specificity, and sensitivity and in **Fig. 7a-b** for mean log-loss score, the sequential classification strategies approached or improved in individual performance measures over fixed strategies that used more data points and/or longer observation intervals.

**Fig. 7:**
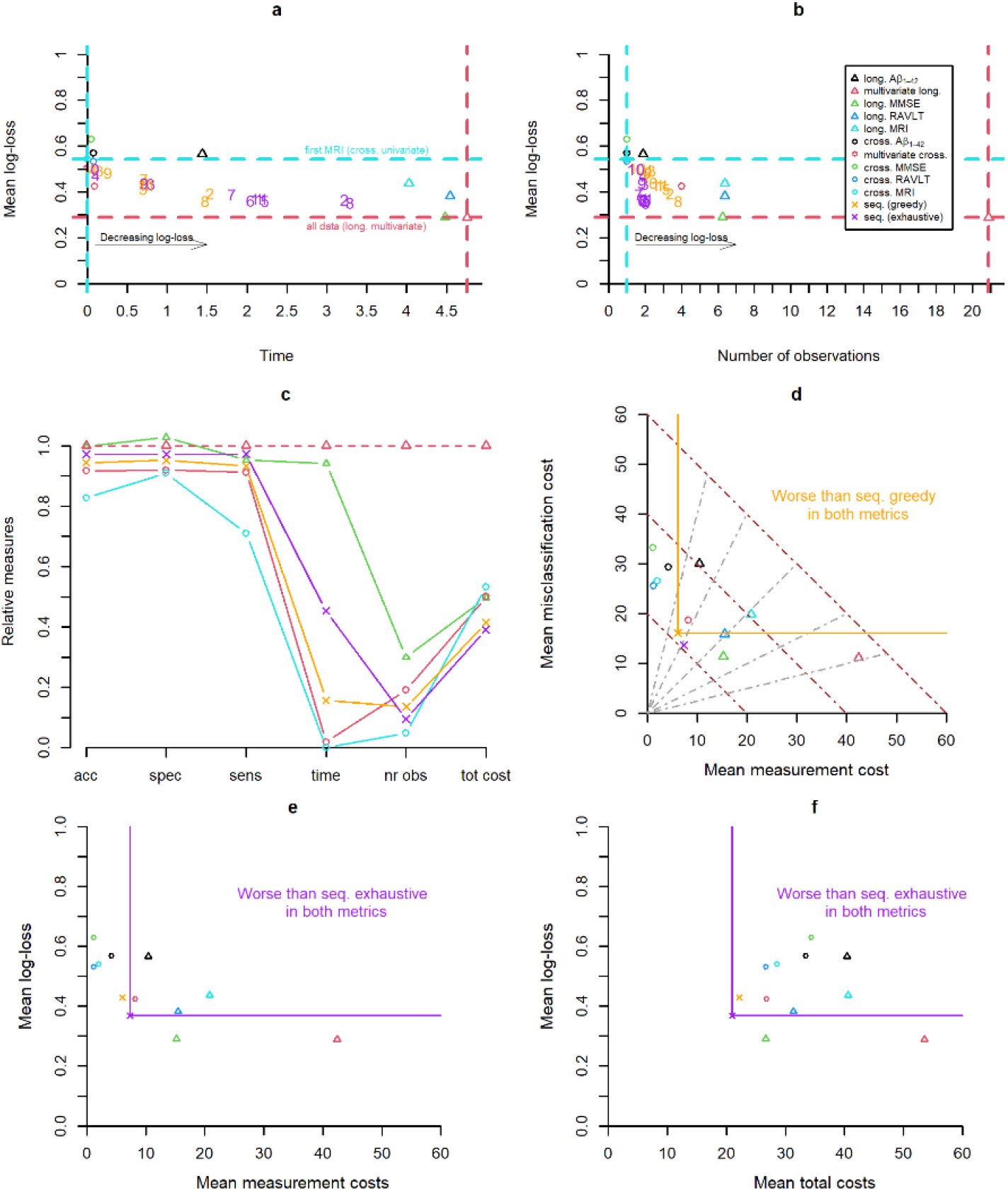
Quantitative comparison of sequential (see prescriptions in caption of Fig. 6) and non-sequential i.e., univariate, or multivariate cross-sectional (cross.) or longitudinal (long.) strategies. **a** Mean follow-up time of a strategy and resulting mean log-loss. **b** Mean number of observations of a strategy and resulting mean log-loss. **c** Strategies in relation to the multivariate longitudinal (for some selected strategies). Performances (accuracy, specificity, and sensitivity) of different strategies divided with the one from the multivariate longitudinal strategy are displayed (represent portion of retained performance). Moreover, the ratios of mean follow-up time or number of observations to the one of the multivariate longitudinal strategy were computed (represent portion of utilized resources). The ratio of the total cost of strategies with the one using all information is also displayed in the right end of the figure (summarizing accuracy and measurement costs of different sources). **d** Two-objective evaluation using the metrics mean misclassification costs and mean measurement costs. Brown dotted lines represent points with same mean total costs and grey dotted lines represent the shares of misclassification and measurement costs from the total costs (middle line: same misclassification and measurement cost, rotated left: higher share of misclassification costs, rotated right: higher share of measurement costs). **e** Two-objective evaluation using the metrics mean log-loss and mean measurement costs. **f** Two-objective evaluation using the metrics mean log-loss and mean total costs.

Moreover, we chose one set of cost parameter to further compare decision strategies with metrics summarizing different sources of costs. From a multi-objective perspective using misclassification costs (equal to the error percentage) and measurement costs as objective metrics, we define that one strategy dominates another when the former undercuts the latter in both metrics. While there was no strategy that dominated all other strategies, sequential strategies dominated more competing strategies than all considered fixed strategies. These and all remaining results of this section are based on the following prescribed cost parameters: *Cost of misclassification=100 (for both diagnoses); Cost for MRI acquisition=2; Cost of acquisition of - Aβ*_*1-42-*_ *CSF =4; Cost of acquisition of cognitive test (MMSE or RAVLT) =1; Cost for waiting one year=2*. We chose relatively small costs of delaying and acquisition to encourage an accuracy close to the maximum but with fewer measurements and shorter follow-up time. In **Fig. 7c** a subset of decision strategies was evaluated relative to the idealized performance of using all available information. The univariate cross-sectional strategy using the first SPARE-AD performed worst, followed by the multivariate cross-sectional and greedy sequential strategies, the exhaustive sequential strategy and the one using all MMSE measures (showed lower sensitivity but higher specificity). For all considered strategies the relative number of observations and/or the time to diagnosis were substantially reduced. Strategies based on fixed panels that were dominated by a sequential strategy in two metrics are indicated in **Fig. 7d-f**. An evaluation of decision strategies in terms of misclassification, measurement costs and total costs revealed that both sequential strategies showed lower total costs than all fixed, lower misclassification than all cross-sectional, and lower measurement costs than all longitudinal strategies while all univariate cross-sectional strategies had lower measurement costs and some longitudinal strategies lower misclassification costs (see **Fig. 7d**). As visualized in **Fig. 7d** the multivariate cross-sectional, and univariate longitudinal strategies using all SPARE-AD or all A*β*_1-42-_ CSF measures performed worse than the greedy sequential strategy in both misclassification and measurement costs. The exhaustive sequential strategy dominated additionally also the univariate longitudinal strategy using all RAVLT measures. **Figure 7e-f** visualizes for the exhaustive strategy the region in which strategies are dominated in two objectives, either the mean log-loss and the mean measurement costs or the mean log-loss and the mean total costs.

### Sensitivity in predicting conversion before clinical manifestation

The results of longitudinal strategies so far covered metrics that did not consider whether the prognosis of conversion was concluded before or after the conversion to manifest AD occurred. To address this aspect, we considered an objective metric assessing the suitability of disease prognosis by counting correct diagnoses after the conversion to AD as an error. We defined the pre-conversion sensitivity as the portion of MCI-converters that were correctly classified before the conversion occurred. Increasing the costs of time led to lower follow-up times of sequential strategies and consequently higher pre-conversion sensitivities (**Fig. 8a**). As expected, fixed longitudinal strategies had pre-conversion sensitivities of around 0 and for cross-sectional strategies the pre-conversion sensitivity was equal to the sensitivity (**Fig. 8b**). As displayed in **Fig. 8b**, the portion of retained sensitivity (pre-conversion sensitivity divided by sensitivity) scatter between one fourth (25% of all correctly predicted conversions were made in the MCI-state) and one (all correctly predicted conversion were made before clinical manifestation). **Figure 8b-d** illustrates that there is a trade-off between pre-conversion sensitivity on the one side and accuracy, specificity, and sensitivity on the other side as strategies with high pre-conversion sensitivity tend to be less accurate, specific and sensitive. The performance of sequential strategies was upper-bounded by the pre-conversion sensitivity of multivariate cross-sectional strategy and the accuracy of the multivariate longitudinal strategy. Strategies with low mean follow-up time showed similar pre-conversion sensitivity, accuracy and specificity as the multivariate cross-sectional strategy. The sequential strategy with highest pre-conversion sensitivity of 0.82 achieved a specificity of 0.79 and accuracy of 0.80 using in average 1.9 measurements assessed over a time interval of around 40 days. This sequential strategy achieved 99 percent of the highest pre-conversion sensitivity of the cross-sectional multivariate strategy (with 99 percent of accuracy) that included four measurements for every subject and a mean follow-up time of 34 days. With increasing mean follow-up times, strategies tended to be more accurate and specific while the pre-conversion sensitivity dropped to 0.18. Greedy sequential strategies tended to have higher pre-conversion sensitivities than exhaustive sequential strategies while being less accurate.

**Fig. 8:**
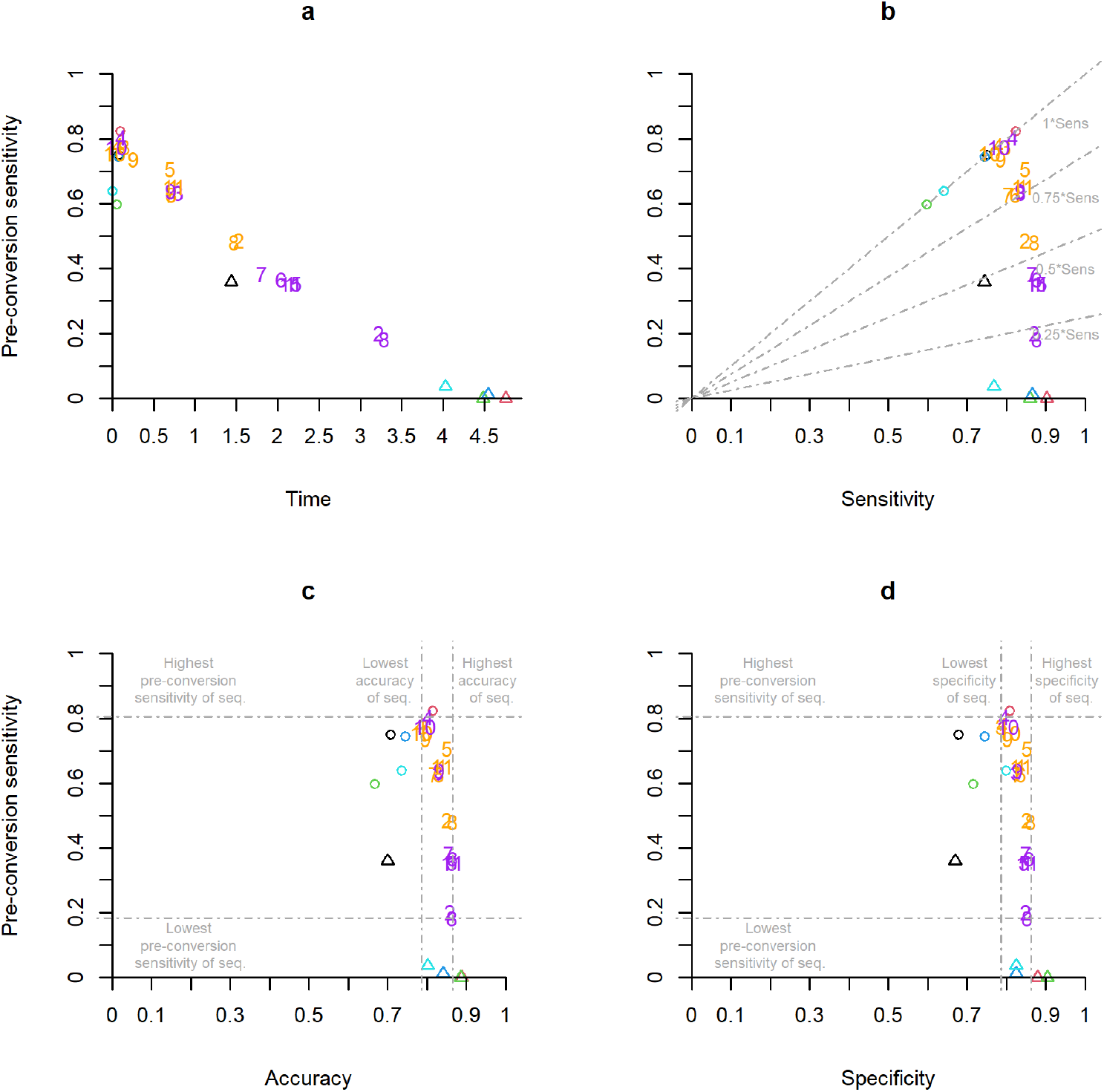
Comparison of decision strategies (see caption of Fig. 6 for the prescription of cost parameters for the sequential strategies) according to their pre-conversion sensitivity and corresponding **a** mean follow-up time, **b** sensitivity (grey dashed lines indicate lines where the pre-conversion sensitivity is equal to portions of either 1, 0.75, 0.5 or 0.25 of sensitivity), **c** accuracy **d** specificity.

## Discussion

Personalized medical care tailors the diagnosis and treatment to individual patients based on circumstantial evidence and options.^15^ Motivated by a use case of pre-clinical diagnosis of AD dementia —the most frequent cause of severe cognitive decline at old age, we developed a general statistical framework to substantiate the selection of sequence and time of diagnostic markers to include in a decision process. As previous studies indicated a benefit of combining multiple, diverse markers^6^ we included cognitive, brain anatomical, and molecular markers. The accuracy of the non-sequential strategies consistently improved when multiple cross-sectional and/or multiple longitudinal measurements were combined. This confirmed that the implemented longitudinal discriminant model was able to leverage complementary information across markers and time contained in multi-variate repeated, irregularly sampled measurements. The observed complementary information of different diagnostic markers has been shown and explored in the literature previously^6^ and the performance achieved with all available data marks the maximum accuracy that can be expected from sequential algorithm with fewer variables or time points. However, under certain conditions, a single or very few markers may be sufficient for a confident diagnosis, and it is the objective of sequential neutral zone classifiers to balance misclassification and measurement costs. By prescribing application-dependent cost parameters, we tuned the trade-off of accuracy, number of measurements, and delay of diagnosis. The results of the sequential classification strategies showed the expected benefits that comparable accuracies could be reached with fewer acquisitions and shorter follow-up interval.

We demonstrated how accuracy and the fraction of patients receiving an optional invasive molecular assessment alongside a regular MRI was calibrated by the prescribed measurement costs of A*β*_1−42_ - CSF. The inclusion of more invasive measures led to higher sensitivity at almost constant specificity. The prescription of optimal measurement costs depends on the specific use case and was not addressed in the scope of this work. Using only MRI, most false negatives as well as false positives were A+T+. When A*β*_1−42_ -CSF was included, more MCI-converters with A+T+ were correctly classified. On the other hand, A+T+ MCI-stables were more frequently falsely classified when A*β*_1−42_ -CSF was included, while for profiles with A-less false positive classification were made. In about half of falsely positive predicted MCI-stables with both cross-sectional biomarkers, considering all cognitive measures for prediction led to correct classifications. According to our model, the biological and cognitive measurements of these cases together make a conversion within three years less likely even though the cross-sectional biomarkers alone indicate a future conversion. Hence, they might show AD pathology without the cognitive progression to dementia. Some MCI-stables were even falsely classified when using MRI, A*β*_1−42_ -CSF and all cognitive measures. In an extreme example, the participant had an MMSE score of 16 and otherwise indicators of AD, but was recorded in the ADNI data base as not having AD. While a clerical mistake cannot be excluded, we note that the labels of the AD spectrum are uncertain and the manifestation of the disease effect on brain anatomical structure^16,17^, molecular^18^, and cognitive profile^19^ is heterogeneous.

In the second application we included longitudinal measurements of biological and cognitive markers for sequential classification. We calibrated the accuracy-costs trade-off by varying cost parameters. When taking more observations (by lowering costs of marker acquisition) or waiting for a longer time (by lowering costs of time) accuracy of sequential strategies approached the highest accuracy achieved when combining all available data of the subjects. When increasing costs of time, conversion was predicted before progression to manifest AD more often, making the setting better suited for prognosis but at the cost of both specificity and sensitivity. Interestingly, higher costs of acquisition did not result in a drop of accuracy but in a drop of pre-manifest sensitivity. The implemented greedy sequential strategies based on high acquisition costs considerably reduced the number of observations (especially of A*β*_1−42_ -CSF) and instead chose to assess the cognitive MMSE scale after a longer follow-up time when gains in accuracy pay out against the high acquisition costs. For the greedy sequential strategy with default cost parameters the waiting time until a decision was 0.74 years in average, whereas 30 percent of the participants were definitively classified based on a single measurement. On average, 2.8 measurements were used to conclude the prognosis. Only 14 percent of all available measurements were selected at the cost of specificity (−0.04) and sensitivity (−0.06). Since on average the last measurement was assessed after 0.74 years--around 4 years earlier than when using all measurements, the strategy could also detect future conversions before clinical manifestation (sensitivity of 0.84 and pre-manifest sensitivity of 0.65). All these considerations are relevant when aiming to prescribe cost parameters in clinical diagnosis. We chose one set of cost parameters to examine potential benefits of sequential classifiers over classifiers based on fixed panels of measurements. Multi-objective evaluation for a given cost prescription revealed that sequential strategies could also dominate other non-sequential strategies by making less errors and at the same time causing less measurement costs. Individualized measurement sequences undercut panels containing all cross-sectional multivariate data or longitudinal measurements of one biomarker (MRI or A*β*_1−42_ -CSF) in both objectives for the considered prescription of cost parameters while no competing non-sequential strategy dominated the sequential strategies.

Mixed-effects models were used in earlier studies to model univariate or multivariate repeated (longitudinal) clinical data.^20–31^ Because of their ability to integrate irregular sampling intervals and varying sequence lengths, mixed-effects models were applied for medical diagnosis with longitudinal data in general ^32–41^ and in the field of neurodegeneration in particular^23,38,41^. In recent years, mixed-effects models were implemented to derive flexible predictions based on variable subsets of measurements or dynamically updating predictions in case new measurements were collected.^23,25,28,36,42^ Mixed-effects model based empirical Bayes classifiers allow to fit a single model that is then used for varying predictive clinical applications, a property that might be important when it comes to the translation into clinical practice. Rather than training multiple models for a wide range of plausible diagnostic scenario, our generative approach does not require re-training of the distribution parameters for new applications as long as the variables are the same. In the field of neutral zone prediction, multiple approaches were presented in the last years.^7–9,42,43^ The existing prospective sequential neutral zone classifiers were designed for multi-stage classification which is limited to the choice of whether to include another marker.^7,8,42^ In contrast, our more flexible algorithm can skip observations and can choose which type of marker to select next. Due to the ability of mixed-effects models to adapt model structures to the data, task, and sample size, our framework of non-sequential and sequential classification based on mixed-effects models may be of interest in different applications and research fields. Our computational framework is publicly available as an R package. The specific implementation presented in this study is limited to logistic regression for prevalence and linear mixed-effects models for modelling the marker distributions. Nevertheless, the concept is applicable to other modelling approaches (e.g., non-linear mixed-effects models) that deliver the estimated distribution parameters (prevalence, population means and covariance matrices) needed for the application of decision and selection rules.

While the methodology is generally applicable to a variety of tasks, the evaluation of the application in this piece has some limitation. The classification task was defined by clinically motivated, yet arbitrary, thresholds of follow-up and conversion time. Moreover, in the two studies, participants with significant neurological disorders and most psychiatric disorders were excluded, limiting the validity of an application to a prospective clinical population as shown previously in applications of machine-learning methods to data from clinical routine.^44,45^ Here we evaluated fixed cost parameters. In a clinical application, the costs could depend on the visit or on other factors. For instance, as already implemented in many clinical workups, initial suspicion of dementia due to AD requires a confirmatory MRI to exclude other neurological disorders. In our framework, this would lead to a cost penalty of zero in case of a suspected case of AD or worded differently: “no definitive diagnosis of AD without structural MRI”. The simplified estimation of the distribution underlying the predictive model may limit the performance of the model and its application to clinical populations. Each marker and random effect of each marker increases the dimensionality of the covariance matrix, thereby setting limits on how complex fitted models can be before the numerical estimation of the covariance matrix parameters does not converge anymore. In our experiments, convergence 5000 or fewer iterations, depending on the fold. Notably, the models converged on multiple systems (Linux and Windows) but with slightly different model parameter estimates and different number of iterations required to reach convergence. Fitting models with more variables without more observations could be achieved by fitting all pairwise LMMs covering the data of only two variables while averaging estimates that are trained multiple times.^22^ While effective and computationally light, we implemented an approach selecting a single measurement in a sequence which does not guarantee to find the globally optimal next step. By adapting the multi-stage approach presented in^7^ in a way that allows to choose which observation should be taken next, better sequential classification might be possible. A globally optimal algorithm would need to consider all possible future sequences which is computationally intensive and intractable for large numbers of markers.

Academic research of biological or behavioral markers is often focused on methods to build good predictive model with a single or a few general-purpose modalities.^46^ Acquisition of a complete set of markers for all participants with high accuracy on population-level might still be insufficiently certain for clinical diagnosis in certain individuals. It is relevant to know which diagnoses are uncertain to achieve individualized precision diagnostics. An individualized sequence of markers as presented here, follows the principle of precision diagnostic of “use it only when and in whom it is most needed”. While the proposed sequential algorithm does not alleviate the necessity of choosing cost parameters, it demonstrated the ability to adapt out-of-sample classification to consistently lower costs compared to cross-sectional predictions or the ones using all available information of one or multiple markers we have for a subject. Sequential strategies that were designed to lower overall decision costs showed to be competitive in performance measures using less resources. The proposed statistical framework constitutes a promising element for precision diagnostic that makes the panel of diagnostic markers conditional on past and potential future evidence, thus specifically individualizing the acquisition of the panel of markers after each visit. Despite the presented methodological strengths and potential benefits, the implementation into consequential clinical workups is not supported by our findings and up for debate. The outcome—even when neglecting potential biases and uncertainty, intrinsically depends on inherently subjective prescribed cost parameters. These parameters express a variety of multi-faceted quantities such as monetary acquisition cost, physical and psychological patient burden, time-to-decision, and many others in a single unit. Only if the range of prescribed costs is widely agreed upon, and one method dominates another across the entire range, then superiority can be claimed.

## Materials and Methods

### Decision rules for classification

Suppose we have a class label *z* ∈ {1; 2}, a random binary variable with Bernoulli distribution that has an unconditional success probability π_0_ (prevalence) and ***y*** ∈ ℝ^*m*^, a random vector taking continuous values that has a density *ϕ*^(1)^ for the population assigned class *z* = 1, and density *ϕ*^(2)^ for the population assigned class *z* = 2. In this section we derive the decision rules for a multi-variate normal distribution with equal covariance in each class. The more general case of unequal covariance across classes is described in the Supplementary Methods 2. We assume that:

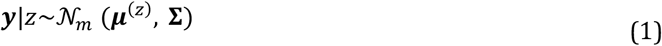

with mean vectors ***μ***^(*z*)^ = *E*(***y***|*z*) (*z* ∈ {1; 2}, ***μ***^(1)^ ≠ ***μ***^(2)^) and the common covariance matrix **Σ** = **Σ**^(1)^ = **Σ**^(2)^ = Var(***y***|*z*). The posterior probability of *z* = 2 given ***y*** is:

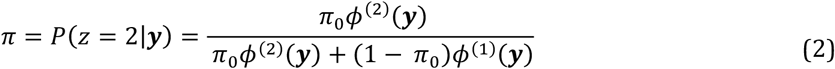

Suppose 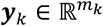 are past observations at present time, 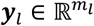 are optional future observations, and, without loss of generality, let 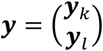. The posterior probability at present *π*_*k*_ is given by the observed values ***y***_*k*_ and we call it current evidence as it incorporates the entirety of made observations up to present. Based on the current evidence, a forced choice classifier δ_*F,C,k*_ definitively assigns one of two classes in a binary classification. We denote with 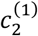 the costs of a false positive and 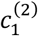 the costs of a false negative classification. The decision rule of the forced choice classifier δ_*F,C,k*_ that minimizes misclassification costs and the corresponding expected misclassification costs *EC*_*k*_ after ***y***_*k*_ was assessed are

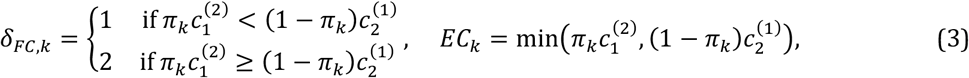

Neutral zone classifiers add a no decision class *NZ* (neutral zone) to the set of possible predicted outcomes.^9,42,43^ We derived such classifiers by prescribing additional measurement costs *c*^*ℳ*^. These costs account for postponing the decision and acquiring additional markers ***y***_*l*_. A definite decision is postponed only if the expected future costs given by a classification with ***y***_*l*_ conditional on the value of the already assessed ***y***_*k*_ denoted by *EC*_*l,k*_ are lower than *EC*_*k*_ (based on assessed ***y***_*k*_). The costs *EC*_*l,k*_ are the sum of the measurement costs and the expected future misclassification costs. Consequently, for *EC*_*l,k*_, the expected false positive and false negative rate of a forced choice classification based on the joint distribution of ***y***_*l*_ and *z* conditional on ***y***_*k*_ is needed. In linear discriminant models, both misclassification rates depend solely on the misclassification costs 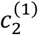 and 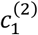, current evidence *π*_*k*_ and the prospective discriminability denoted by Δ_*l,k*_ (see Supplementary equation S15). It is defined as 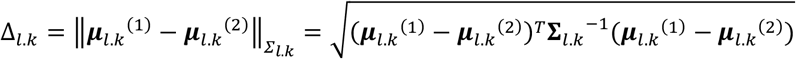 where ***μ***_*l,k*_^(*z*)^ and **Σ**_*l,k*_ are the parameters of the distributions 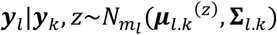 (given in Supplementary Equation S1). The prospective discriminability quantifies the expected added discriminative value of including ***y***_*l*_. We denote the false positive rate by *FP* (*π*_*k*_, Δ_*l,k*_) and the false negative rate by *FN* (*π*_*k*_, Δ_*l,k*_) for a classification based on the joint distribution ***y***_*l*_, *z*|***y***_*k*_ (see Supplementary Equation S15 for the closed form solution). The expected costs *EC*_*l,k*_ is

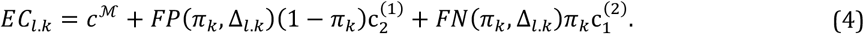

Given the expected costs we can derive a neutral zone classifier that assigns the label *NZ* whenever *EC*_*k*_ > *EC*_*l,k*_. In case *EC*_*k*_ ≠ *EC*_*l,k*_ the prospective neutral zone classifier δ_*PNZ,K*_ is given by:

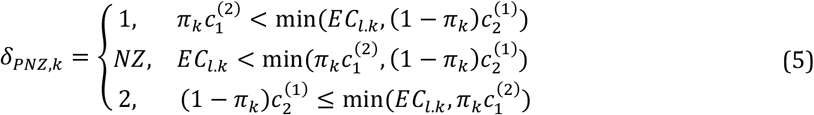

In this study we set both misclassification costs to 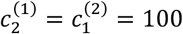 such that the measurement costs *c*^*ℳ*^ can be interpreted as the percentage of one misclassification. Hence, if one measurement costs *c*^*ℳ*^ then the classifier δ_*PNZ,K*_ predict the label *NZ* whenever the expected increase in accuracy by including ***y*** is higher than 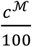 (e.g. *c*^*ℳ*^=4: classifier predict NZ if the expected accuracy increase is higher than 0.04). More information and the derivation can be found in the Supplementary Methods 2 in general and Supplementary Equations (S11) and (S12).

### A multi-variate longitudinal discriminant model for sequential classification

In this study mixed-effects model-based estimation was embedded into linear (or quadratic, see Supplementary Methods 3-4) discriminant models to account for inter-subject differences ^32–35,40,41,47^ (**Fig. 9A-B**). The vector ***y***_*i*_ from participant *i* ∈ {1; 2; … ; *n*} consists of longitudinal measurements *y*_*i,j*_ from multiple time points *t*_*i,j*_ acquiring one of four markers (*H* denoting the set of names of all considered markers). We assumed for a subject *i* with measurements ***y***_*i*_ and unknown label *z*_*i*_ that

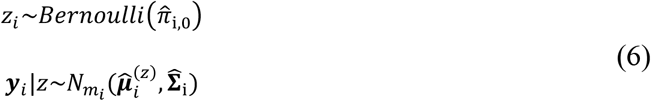

**Fig. 9.**
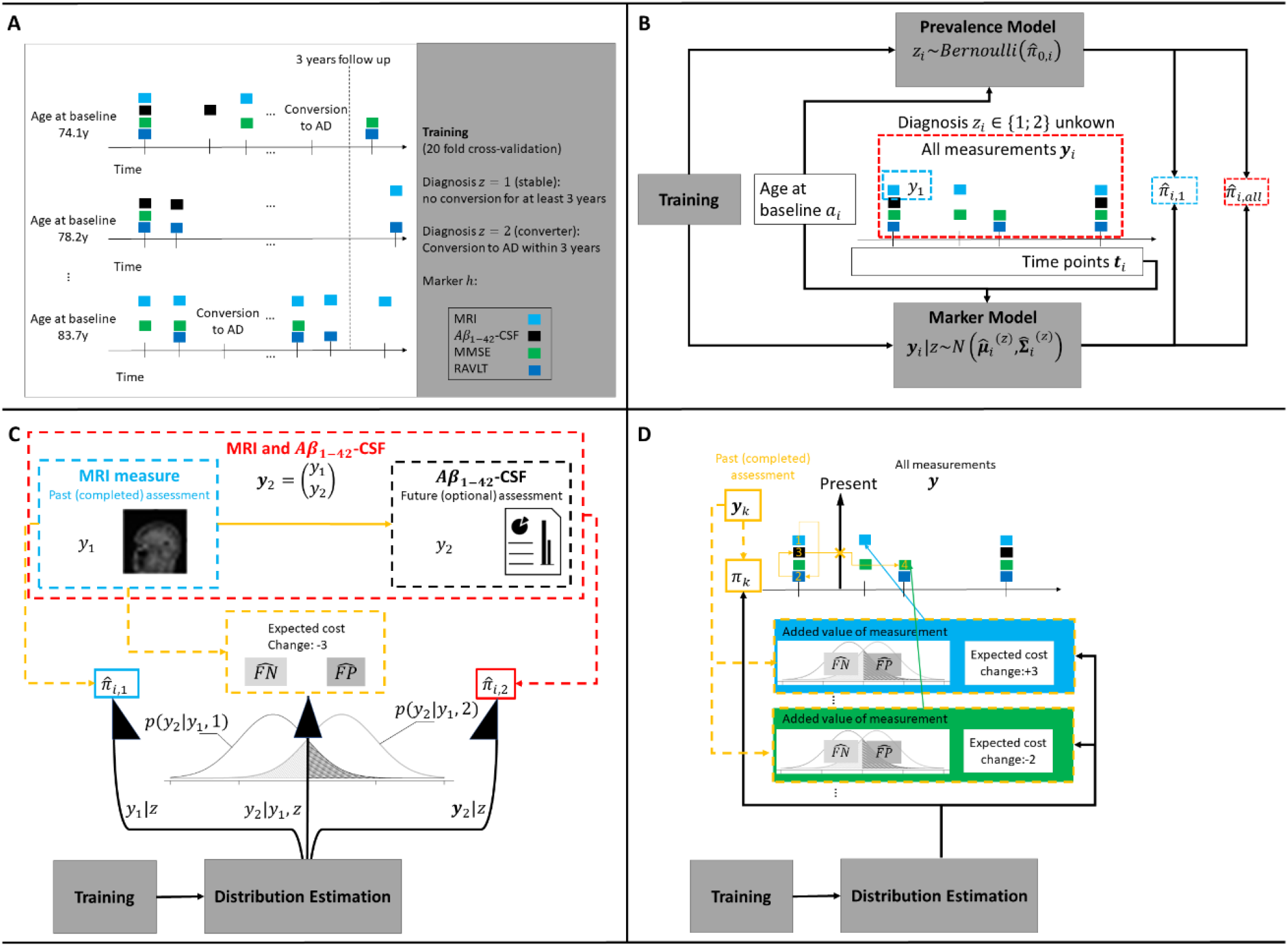
Illustration of the proposed classification framework. (a) Training: Linear mixed-effects models were trained (20 fold cross-validation) on irregular, multi-variate longitudinal data to derive classifiers separating patients with mild cognitive impairment that either converted to AD within three years or less, or stayed stable for three years or more. (b) Inference: Distributions (means and covariances) of the markers (MRI, *Aβ*_1−42_ -CSF, MMSE or RAVLT) were estimated using the age at baseline and the time points at which the observations occurred (time since baseline) as predictors. With the estimated distributions and the values of the considered observations, estimators for the posterior probability of being a MCI-converter of arbitrary subsets can be computed (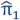 only with first MRI or 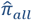 with all observations as examples). (c) Sequential two-stage classifier: Two-stage classifier making classification either with a MRI measure at baseline (posterior probability 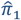) or optionally with both MRI and *Aβ*_1−42_ -CSF measures at baseline (posterior probability 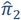). For the decision if the optional measurement is included 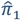 and the estimated prospective misclassification rates (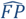 and 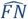) are used to quantify the change in the expected costs. (d) Sequential classifier for longitudinal sequences: Sequential classifier that decide at every step for one of the diagnoses or to postpone the decision and collect more measurements (decision rules) and selects the next observation for classification (selection rule). For the decision at a step *k* after the measurements *y*_*k*_ are assessed, the current evidence 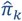 and prospective misclassification rates of leftover measurements are considered.

The prevalence 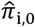 was either estimated using a logistic regression with the age at baseline *a*_*i*_ of a subject as predictor and its diagnosis *z*_*i*_ as response (conditional prevalence) or with the relative frequency of subjects with *z*_*i*_ =2 in the training set 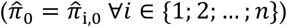. The logistic regression considered a (fixed) intercept *λ*_0_ and (fixed) slope in the baseline age *λ*_0_ as model parameters and we denote with ***λ*** the vector containing both these two parameters (see Supplementary equation S21 for the model equation of the logistic regression). In the main text of the study, we report only results based on models with conditional prevalence. The estimators. 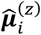 and 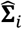 were derived using LMM. The LMM included labelled marker values of observation *j* of subject *i* denoted by 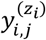 as response and the known diagnosis *z*, the age at baseline *a*, time since baseline *t*_*i,j*_ and four dummy variables for coding the type of marker *ν*_*h,i,j*_ *h* ∈ *H*) as predictors. We used a model with the model equation ^20,30,31^

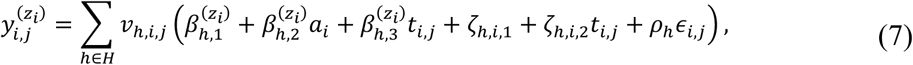

whereas 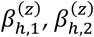 and 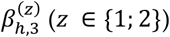 were the diagnosis and marker specific fixed effects and ***β***^(*z*)^ the vector containing all diagnosis-specific fixed effects (population-level), ζ_*h,i*,1_ and ζ_*h,i*,2_ the marker specific random effects (subject-level, same for both labels) and *∈*_*i,j*_ the (scaled) residuals which are multiplied with the marker specific intra-subject variance components *ρ*_*h*_ (same for both labels). The distribution of the vector **ζ**_*i*_ containing all random effects (for the intercept and time for all variables) is given by **ζ**_*i*_ ∼*N*_8_ (0, **Ψ**). The scaled residuals were assumed to be independent from each other and the random intercept and slopes and standard normal distributed i.e.,*∈*_*i,j*_ ∼ *N*(0,1). The distribution of the unscaled residuals ε_*i,j*_ varies between markers and is given as *ε*_*i,j*_ ∼*N*(0, **Σ**_*h*∈*H*_ *ν*_*h,i,j*_ ρ_*h*_), We denote with ***ρ*** = (*ρ*_*h*_)_*h*∈H_ the vector containing all marker-type-specific intra-subject variances. All parameter ***θ*** = **[*λ***; ***β***^(1)^; ***β***^(1)^; **Ψ**; ***ρ***] (respectively ***θ*** = [*π*_0_ ; ***β***^(1)^; ***β***^(1)^; **Ψ**; ***ρ*]** in case of constant prevalences) necessary to specify the model in Equation 6 were estimated on training data using a 20-fold cross validation framework. The estimation of the parameters of the LMMs were obtained with the restricted maximum likelihood approach.^48^

With the estimated prevalence, mean vectors, and covariance matrix we computed all previously derived quantities (posterior probabilities, expected misclassification rates, expected costs and classifiers) by plugging in 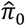 and components of 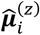 and 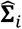 of subject *i* for the true population values in the equations. Moreover, we derived a sequential classification approach that stepwise adds new observations. First, we derived a sequential two-stage neutral zone classifier using the data of cross-sectional biomarkers (**Fig. 9C**). The classifier uses the MRI measurement to either classify the subject as stable, converter, or neutral (*NZ*). In case the label *NZ* was assigned, an A*β*_1−42_ -CSF was added to conclude the prognosis with a forced-choice. This classifier is a special version of the more general multi-stage classifier derived in an earlier study.^7^ As second application, we derived a sequential neutral zone classifier for longitudinal data with the ability to skip inclusion of markers (**Fig. 9D**). The sequential classifier definitively predicts for a subject *i* at the step *k* (1 ≤ *k* ≤ *m*_*i*_) one of the possible prognoses or makes no decision when the prediction falls into the neutral zone. Following, a non-definitive prognosis, the algorithm employs its selection rule to decide which future measurement should be included next. The proposed sequential decision rule (see Supplementary Equation (S24)) applied the prospective neutral zone classifier formalized in Equation 6 to all future observations separately. We can think of applying the prospective neutral zone classifier for every left-over observation separately and assigning the label *NZ* if at least for one observation the prospective neutral zone classifier reveals the label *NZ* as outcome. In case the label *NZ* was chosen a selection rule is applied to choose which (single) observation is included next for the prediction. Two alternative selection rules were evaluated. The greedy rule selected the earliest observation with expected cost reduction and the exhaustive rule selected the observation with highest expected cost reduction.

### Statistical software implementation

Our framework for **P**r**O**spective **SE**quent**I**al **D**iagn**O**sis with **N**eutral zones (POSEIDON) based on estimates from multivariate linear mixed-effects classification models is implemented in the statistical programming language R^49^ (version 4.1.2) and available at https://git.upd.unibe.ch/openscience/POSEIDON. The library depends on the packages *nlme*^48^, *emdbook*^*50*^ and *mixedup* (description and availability at https://github.com/m-clark/mixedup). The POSEIDON library includes functions to train multi-variate linear-mixed effects classification models, make inference with individual sequences of participants and select subsets of all available measurement sequentially for classification. These three steps are combined to derive individualized panels for one participant. Documentation and a full example with synthetic data is included in the repository. The example uses a synthetic data set to train a model on the data of all subjects except one and the make (out-of-sample) inference and classifications for the remaining subject. The synthetic data set consists of marker values simulated from a classification model trained with POSEIDON using the age at baseline, time points of the measurement and diagnosis of the subjects considered in this study.

The prevalence is either modelled as constant or as a function of subject characteristics by fitting a logistic regression. The LMMs for modelling the repeated measurements were models constrained to one upper-level clustering variable (e.g., repeated observations within subjects) but arbitrary fixed and random effect’s structures can be specified for such a nesting structure. For our applications with longitudinal data, we assumed the model structure as in Equation 7 for linear, and Supplementary Equation (S20) for quadratic discriminant models. More detailed model descriptions of LMMs in the context of classification (including the heterogeneous case) and the modelling of longitudinal data are provided in the Supplementary Methods 3.

### Study data and empirical evaluation

Longitudinal data from individuals from Alzheimer’s Disease Neuroimaging Initiative (ADNI)^51^ and the Australian Imaging Biomarkers and Lifestyle flagship study of ageing (AIBL)^52^ were included. AIBL study methodology has been reported previously.^52^ The ADNI was launched in 2003 with the primary goal of testing whether serial magnetic resonance imaging (MRI), positron emission tomography (PET), other biological markers, and clinical and neuropsychological assessment can be combined to measure the progression of mild cognitive impairment (MCI) and early Alzheimer’s disease (AD). For up-to-date information and data access see https://www.adni-info.org and https://adni.loni.usc.edu.

We included biological as well as cognitive markers to separate patients with MCI that do not convert to AD over a follow-up time of at least 2.75 years (MCI-stable, label *z* = 1) or convert to manifest AD within 3.25 years since study entry (MCI-converter, *z* = 2). As structural biomarker we used the SPARE-AD score^13^ computed from regional brain volumes obtained from standard structural MRI with a multi-atlas segmentation algorithm^53^ that captures how “AD-like” the structure of the brain of a participant is. The SPARE-AD score is the average decision value of an ensemble of linear support vector machine classifiers trained to discriminate cognitively healthy subjects from patients suffering from AD based on regional anatomical brain volumes. The ensemble was trained on multi-centric data from the iSTAGING Study after statistical harmonization of the 145 regional volumes using Combat-GAM^54^ that accounted for biological variance due to age, sex, and intra-cranial volume. We include A*β*_1−42_ levels in the CSF ^10^ as invasive, AD-specific marker. Cognitive markers were scores given by either the MMSE or RAVLT. We transformed the MMSE scores using normalization proposed in ^55^. The normalization corrects the poor properties (ceiling/floor effects, varying sensitivity to change) of the raw MMSE such that the normalized scores can properly be assessed with linear mixed-effects model.^55^ The normalization was clinically evaluated covering data of normal or pathological aging. All four markers were irregularly measured with differing time points and number of observations between subjects. Prior to model training we scaled the markers by subtracting the empirical mean and divide by the standard deviations (SDs) of all subject-wise means of all observations of a marker. The means and SDs of all variables were estimated within the 20 fold cross-validation framework, i.e., a subject was scaled by the means and SDs estimated on the data of the 19 folds that did not include the subject. We fitted linear mixed-effects models and included only subjects that match the definition either of MCI-stable or MCI-converter and had at least 8 observations independently of the variable. Eventually, a sample with 612 subjects (343 MCI-stables and 270 MCI-converters) were used to fit the 20 models for cross-validated predictions. For the evaluation of the first application covering two-stage classifier we only included the data of 409 subjects that had at least one MRI followed by one A*β*_1−42_ -CSF within 3 months for evaluation (243 MCI-stables and 166 MCI-converters). For the comparison of the decision strategies of the second application covering diagnosis based on multi-variate, longitudinal measurements, we only used the data of 402 subjects that had at least one observation of each variable within 3 months after the first observation (239 MCI-stables and 163 MCI-converters). More information about the selection of the training and evaluation data sets can be found in the Supplementary **Fig. S1**.

## Data Availability

All data produced in the present study are available upon reasonable request to the authors

## Data availability

Raw imaging data and cognitive scores used for this study were provided from ADNI and AIBL studies via data sharing agreements that did not include permission to further share the data. Data from ADNI and AIBL are available through the LONI database (adni.loni.usc.edu) upon registration and compliance with the data usage agreement for each study separately.

## Code availability

The core functions and a full example including data to fit a model and apply it to unseen data are available as R package at https://git.upd.unibe.ch/openscience/POSEIDON. Additional code used to e.g., create figures or tables will be shared upon reasonable request.

## Acknowledgments

The iSTAGING study is a multi-institutional effort funded by NIA through RF1 742 AG054409. AA was funded through grants 191026 and 206795 awarded by the Swiss National Science Foundation. ADNI (National Institutes of Health Grant U01 AG024904) and DOD ADNI (Department of Defense award number W81XWH-12-2-0012) are funded by the National Institute on Aging, the National Institute of Biomedical Imaging and Bioengineering, and through generous contributions from the following: AbbVie, Alzheimer’s Association; Alzheimer’s Drug Discovery Foundation; Araclon Biotech; BioClinica, Inc.; Biogen; Bristol-Myers Squibb Company; CereSpir, Inc.; Cogstate; Eisai Inc.; Elan Pharmaceuticals, Inc.; Eli Lilly and Company; EuroImmun; F. Hoffmann-La Roche Ltd and its affiliated company Genentech, Inc.; Fujirebio; GE Healthcare; IXICO Ltd.; Janssen Alzheimer Immunotherapy Research & Development, LLC.; Johnson & Johnson Pharmaceutical Research & Development LLC.; Lumosity; Lundbeck; Merck & Co., Inc.; Meso Scale Diagnostics, LLC.; NeuroRx Research; Neurotrack Technologies; Novartis Pharmaceuticals Corporation; Pfizer Inc.; Piramal Imaging; Servier; Takeda Pharmaceutical Company; and Transition Therapeutics. The Canadian Institutes of Health Research is providing funds to support ADNI clinical sites in Canada. Private sector contributions are facilitated by the Foundation for the National Institutes of Health (www.fnih.org). The grantee organization is the Northern California Institute for Research and Education, and the study is coordinated by the Alzheimer’s Therapeutic Research Institute at the University of Southern California. ADNI data are disseminated by the Laboratory for Neuro Imaging at the University of Southern California.

## Author contributions

P.W., D.G., S.K., and A.A. designed the research; P.W., D.G., and A.A. performed research; P.W. implemented the algorithms; P.W., D.G., H.S., C.D., S.K., and A.A. analyzed and interpreted the data; P.W., D.G., H.S., C.D., S.K., and A.A. wrote the paper.

