## Supplementary Materials for "Adaptive data-driven selection of sequences of biological and cognitive markers in pre-clinical diagnosis of dementia"

### Supplementary Methods 1: Costs of decision processes

A decision process $d$ incorporates a subset of all available data for its decision according to a decision rule and—in case of sequential classifiers, also a sequence selection strategy. The cost of the decision process is an empirical metric of performance (lower is better) that is the sum of multiple costs each reflecting a different desirable quality. We implement binary decision processes that predict whether a participant $i \in \{1, . . , n\}$ will progress from a sub-clinical stage of mild cognitive impairment (MCI) to the manifest stage of dementia due to Alzheimer's disease (AD) within a time interval of approximately three years. The total costs $c_{d,i}^{\mathcal{T}}$ of a process p are defined as the sum of misclassification costs $c_{d,i}^{\mathcal{C}}$ and measurement costs $c_{d,i}^{\mathcal{M}}$ consisting of the costs of delaying the decision $c_{d,i}^{\mathcal{D}}$ and costs of acquisition $c_{d,i}^{\mathcal{A}}$, i.e.

$$c_{d,i}^{\mathcal{T}} = c_{d,i}^{\mathcal{C}} + c_{d,i}^{\mathcal{M}} = c_{d,i}^{\mathcal{C}} + c_{d,i}^{\mathcal{D}} + c_{d,i}^{\mathcal{A}}.$$

Participants that do not progress to manifest AD are assigned the class label $z = 1$; participants that do progress to manifest AD are assigned the class label $z = 2$. The diagnosis is based on one or more optional visits $j \in \{1, \dots, m_i\}$ in which a measurement $y_{i,j}$ assessed at time point $t_{i,j}$ of one of multiple disease markers $h_{i,j} \in H = \{\mathrm{MMSE}, \mathrm{RAVLT}, \mathrm{A\beta_{1-42}}\ \mathrm{CSF}, \mathrm{SPARE-AD}\}$ is taken. The vector $\boldsymbol{y}_i$ contains all available $m_i$ measurements of subject $i$ from all markers over all time points. A process $d$ chooses a sequence of $k_{d,i}$ observations identified by the set of indices $M_{d,i} = \{j_1, j_2, j_3, \dots, j_{k_{d,i}}\} \subseteq \{1, \dots, m_i\}$. Each classification of an individual with the process $d$ carries a cost defined as a weighted sum of cost parameters that characterize various aspects of model performance.

Misclassification costs $c_{d,i}^{\mathcal{C}}$ for participant $i$ depend on the class label and are defined as

$$c_{d,i}^{\mathcal{C}} = \begin{cases} c_2^{(1)} & \text{for classifying 1 as 2,} \\ c_1^{(2)} & \text{for classifying 2 as 1,} \\ 0 & \text{for a correct classification.} \end{cases}$$

The costs for delaying a decision of participant $i$ is proportional to the time lapse $t_{i,j_g} - t_{i,j_{g-1}}$ of the visit $g$ with respect to the last completed visit $g - 1$, irrespective of the conversion status. We prescribe that the cost is $c_t$ per every year of delay, resulting in costs of $c_t\left(t_{i,j_g} - t_{i,j_{g-1}}\right)$ for the time of postponing the decision until visit $g$. The cost of an assessment includes the material cost of the acquisition of a disease marker and the associated patient burden. The cost of acquisition is denoted as $c_{h_{i,j}}$ for $h_{i,j} \in H$. For the whole sequence of measurements of subject $i$, the delaying costs are $c_{d,i}^{\mathcal{D}} = c_t t_{d,i}^{\max}$ with $t_{d,i}^{\max} = \max_{j \in M_{d,i}}\left(t_{ij}\right)$ and the costs of acquisition are $c_{d,i}^{\mathcal{A}} = \sum_{j \in M_{d,i}} c_{h_{i,j}}$.

If the sequence $M_{p,i}$ of a of a process is fixed, it includes all pre-defined assessments independently of the evidence given by the previously assessed measurements. The adaptive strategies sequentially weighing the expected accuracy from acquired evidence against expected gain in accuracy and costs of acquiring new data and delaying the decision.

### Supplementary Methods 2: More information about decision rules of classification

#### Overview

In this section we consider different classifier. First, we derive equations for forced-choice classifiers, i.e., classifiers that assigns one of the possible classes by minimizing the expected misclassification costs. In a next step we introduce a "no-decision" classifier from the family of neutral zone classifier that allows to not choose one of the possible classes by staying in the so-called neutral zone. Such a classifier assumes that subjects can stay in the neutral zone i.e., that we never have to make a definite decision for them because misclassification costs would be too high. They minimize the expected costs for situation where one can dispense to (never) select one of the possible classes for subject with high expected misclassification costs. While they are optimized for non-sequential classification where no definite decision can be made, they were also, implemented within sequential approaches in earlier studies[1] where at the end a forced-choice classification was made. Finally, multiple sub-sections are dedicated for a prospective neutral zone classifier that take into account the added value of optional measurements such that subjects only are assigned to the neutral zone in case expected costs of a forced-choice classification can be reduced when the optional measurements are included as well. These classifiers specifically considers that the label neutral zone can only be assigned temporary. Prospective neutral zone classifier minimize the expected (total) costs for situations where at the end one of the possible classes has to be chosen but optional measurements can be selected conditional on already passed measurements.

As in the main text of this article we consider a random vector $\boldsymbol{y} \in \mathbb{R}^m$ and the class label $z \in \{1; 2\}$ a random Bernoulli-distributed variable with prevalence $\pi_0 = P(\mathrm{z} = 2)$. We assume that $\boldsymbol{y} \in \mathbb{R}^m$conditional on $z$ follow a multivariate normal distribution with densities $\phi^{(z)}$ given by the mean vectors $\boldsymbol{\mu}^{(z)} = E(\boldsymbol{y}|z)$ and covariance matrices $\boldsymbol{\Sigma}^{(z)} = Var(\boldsymbol{y}|z)$ $(z \in \{1; 2\})$. For the derivations of equations for the prospective neutral zone classifier, we consider a participation of the vector of measurements $\boldsymbol{y} \in \mathbb{R}^m$ into two sets with either $m_k$ or $m_l$ $(m = m_k + m_l)$ measurements where we denote the vector of all observations of a set with $\boldsymbol{y}_k \in \mathbb{R}^{m_k}$ and $\boldsymbol{y}_l \in \mathbb{R}^{m_l}$. The vector $\boldsymbol{y}_k$ contains passed, completed measurement and the vector $\boldsymbol{y}_l$ optional measurements that might be assessed in future. We write $\boldsymbol{y} = \begin{pmatrix} \boldsymbol{y}_k \\ \boldsymbol{y}_l \end{pmatrix}$ (without loss of generality) such that $\boldsymbol{\mu}^{(z)} = \begin{pmatrix} \boldsymbol{\mu}_k{}^{(z)} \\ \boldsymbol{\mu}_l{}^{(z)} \end{pmatrix}$ and $\boldsymbol{\Sigma}^{(z)} = \begin{pmatrix} \boldsymbol{\Sigma}_{kk}{}^{(z)} & \boldsymbol{\Sigma}_{kl}{}^{(z)} \\ \boldsymbol{\Sigma}_{lk}{}^{(z)} & \boldsymbol{\Sigma}_{ll}{}^{(z)} \end{pmatrix}$ where $\boldsymbol{y}_k|z \sim N_{m_k}(\boldsymbol{\mu}_k{}^{(z)}, \boldsymbol{\Sigma}_k{}^{(z)})$ and $\boldsymbol{y}_l|z \sim N_{m_l}(\boldsymbol{\mu}_l{}^{(z)}, \boldsymbol{\Sigma}_l{}^{(z)})$ $(z \in \{1; 2\})$. To compute expected future cost reduction by including additional measurements for classification the distribution of $\boldsymbol{y}_l, z\,|\boldsymbol{y}_k$ respectively the distributions of $z|\boldsymbol{y}_k \sim$Bernoulli$(\pi_k)$ (with $\pi_k = P(z = 2|\boldsymbol{y}_k)$)and $\boldsymbol{y}_l|z, \boldsymbol{y}_k \sim N_{m_l}\left(\boldsymbol{\mu}_{l.k}^{(z)}, \boldsymbol{\Sigma}_{l.k}{}^{(z)}\right)$ (with $\boldsymbol{\mu}_{l.k}^{(z)} = E(\boldsymbol{y}_l|z, \boldsymbol{y}_k)$ and $\boldsymbol{\Sigma}_{l.k}^{(z)} = Var(\boldsymbol{y}_l|z, \boldsymbol{y}_k)$ $(z \in \{1; 2\})$) are needed. The distributions $\boldsymbol{y}_l|z, \boldsymbol{y}_k$ are given by (follows from [2]):

$$\begin{aligned} \boldsymbol{y}_l, z\,|\boldsymbol{y}_k &\sim N_{m_l}(\boldsymbol{\mu}_{l.k}{}^{(z)}, \boldsymbol{\Sigma}_{l.k}{}^{(z)}) \\ \boldsymbol{\mu}_{l.k}{}^{(z)} &= \boldsymbol{\mu}_l{}^{(z)} - \boldsymbol{\Sigma}_{lk}{}^{(z)}\left(\boldsymbol{\Sigma}_{kk}{}^{(z)}\right)^{-1}\left(\boldsymbol{y}_k - \boldsymbol{\mu}_k{}^{(z)}\right) \\ \boldsymbol{\Sigma}_{l.k}{}^{(z)} &= \boldsymbol{\Sigma}_{ll}{}^{(z)} - \boldsymbol{\Sigma}_{lk}{}^{(z)}\left(\boldsymbol{\Sigma}_{kk}{}^{(z)}\right)^{-1}\boldsymbol{\Sigma}_{kl}{}^{(z)} \end{aligned} \tag{S1}$$

### Expected misclassification costs of a forced-choice classifier

In the main text of this article, we derived decision rules by directly comparing the expected costs of outcomes of a classifier. In the following we consider a test statistic $T = T(\boldsymbol{y})$ and decision boundary $b$ that define regions of classification outcome. We consider the misclassification costs $c_2^{(1)}$ and $c_1^{(2)}$ (as defined previously) and assume for the moment that there are no measurement costs. When assuming that $T(\boldsymbol{y})$ is a function so that higher values of the test statistic are indicative for the class $z = 2$, then a forced choice classifier $\delta_{FC}$ for z is given as:

$$\delta_{FC} = \begin{cases} 1 & \text{if } T(\boldsymbol{y}) < \text{b} \\ 2 & \text{if } T(\boldsymbol{y}) \geq \text{b} \end{cases} \tag{S2}$$

As test-statistic e.g., the posterior probability $\pi = \text{P}(\text{z} = 2|\boldsymbol{y})$ (see Equation (2) in the main article of this study) can be used for which the decision boundary is $b = \frac{c_2^{(1)}}{c_2^{(1)} + c_1^{(2)}}$.

The costs $C(\boldsymbol{y}, z)$ of the forced choice classifier (random variable) is a function of the random vector $\boldsymbol{y}$ and random variable $z$ and we get the expected costs of a forced choice classifier before assessing $\boldsymbol{y}$ as (assuming no measurement costs, i.e., only the misclassification costs):

$$\begin{aligned} \text{E}\big(C(\boldsymbol{y}, z)\big) &= P(\delta_{FC} = z) \cdot 0 + P(\delta_{FC} = 2, z = 1) \cdot c_2^{(1)} + P(\delta_{FC} = 1, z = 2) \cdot c_1^{(2)} \\ &= P(T(\boldsymbol{y}) \geq b|z = 1)(1 - \pi_0)c_2^{(1)} + P(T(\boldsymbol{y}) < b|z = 2)\pi_0 c_1^{(2)} \end{aligned}$$

where $P(T(\boldsymbol{y}) \geq b|z = 1)$ is the expected false positive and $P(T(\boldsymbol{y}) < b|z = 2)$ the expected false negative rate when classifying $z$ based on $\boldsymbol{y}$ via a test statistic $T(\boldsymbol{y})$. The decision boundary $b$ is chosen such that the expected (misclassification) costs are minimized. The function $T(\boldsymbol{y})$ and the boundary $b$ are given by the misclassification cost parameters $c_2^{(1)}$ and $c_1^{(2)}$ and the joint distribution of $\boldsymbol{y}$ and $z$ respectively the probability $\pi_0 = p(z = 2)$ (prevalence) and the parameters $\boldsymbol{\mu}^{(1)}, \boldsymbol{\mu}^{(2)}, \boldsymbol{\Sigma}^{(1)}$ and $\boldsymbol{\Sigma}^{(2)}$ of the distributions of $\boldsymbol{y}|z$ ($z \in \{1; 2\}$). To denote the underlying distribution used for the classification task, we specify the false positive rate as $FP\big(\pi_0, \boldsymbol{\mu}^{(1)}, \boldsymbol{\mu}^{(2)}, \boldsymbol{\Sigma}^{(1)}, \boldsymbol{\Sigma}^{(2)}\big)$ and false negative rate as $FN\big(\pi_0, \boldsymbol{\mu}^{(1)}, \boldsymbol{\mu}^{(2)}, \boldsymbol{\Sigma}^{(1)}, \boldsymbol{\Sigma}^{(2)}\big)$ as a function of the distributional parameters. We can write the expected misclassification costs $E\big(C(\boldsymbol{y}, z)\big)$ as:

$$E\big(C(\boldsymbol{y}, z)\big) = FP\big(\pi_0, \boldsymbol{\mu}^{(1)}, \boldsymbol{\mu}^{(2)}, \boldsymbol{\Sigma}^{(1)}, \boldsymbol{\Sigma}^{(2)}\big)(1 - \pi_0)c_2^{(1)} + FN\big(\pi_0, \boldsymbol{\mu}^{(1)}, \boldsymbol{\mu}^{(2)}, \boldsymbol{\Sigma}^{(1)}, \boldsymbol{\Sigma}^{(2)}\big)\pi_0 c_1^{(2)} \tag{S3}$$

Of note, $E\big(C(\boldsymbol{y}, z)\big)$ are the expected misclassification costs before knowing any measurement of $\boldsymbol{y}$. For the homogeneous case where both populations have different means $\boldsymbol{\mu}^{(1)} \neq \boldsymbol{\mu}^{(2)}$ but a common covariance matrix $\boldsymbol{\Sigma}^{(1)} = \boldsymbol{\Sigma}^{(2)} = \boldsymbol{\Sigma}$ we derived a closed form solution for both expected misclassification rates (and consequently the expected costs $E\big(C(\boldsymbol{y}, z)\big)$). For the heterogeneous case ($\boldsymbol{\Sigma}^{(1)} \neq \boldsymbol{\Sigma}^{(2)}$) we approximated the expected misclassification rates with Monte Carlo simulations. For a given boundary $b$ of a forced-choice classifier (minimizing the expected costs $E\big(C(\boldsymbol{y}, z)\big)$ in equation

(S3)) the expected misclassification costs in case $\boldsymbol{y}$ is already assessed can be written as (depending on the outcome of $\delta_{FC}$ one of the expected misclassification rate is 0 and the other 1 in Equation (S3)):

$$E(C(\boldsymbol{y}, z)|\boldsymbol{y}) = \begin{cases} \pi c_1^{(2)}, & T(\boldsymbol{y}) < b \\ (1-\pi) c_2^{(1)}, & T(\boldsymbol{y}) \geq b \end{cases} = min(\pi c_1^{(2)}, (1-\pi) c_2^{(1)})$$

In the following we will derive the closed form solutions of the misclassification rates for the homogeneous case. To this end, we consider the test statistic denoted by $s$:

$$s = \frac{\left(\boldsymbol{y} - \boldsymbol{\mu}^{(1,2)}\right)^T \boldsymbol{\Sigma}^{-1}\left(\boldsymbol{\mu}^{(1)} - \boldsymbol{\mu}^{(2)}\right)}{\Delta} \quad \text{(S4)}$$

Where $\boldsymbol{\mu}^{(1,2)} = \frac{\boldsymbol{\mu}^{(1)} - \boldsymbol{\mu}^{(2)}}{2}$ and $\Delta = \left\|\boldsymbol{\mu}^{(2)} - \boldsymbol{\mu}^{(1)}\right\|_{\boldsymbol{\Sigma}} = \sqrt{(\boldsymbol{\mu}^{(1)} - \boldsymbol{\mu}^{(2)})^T \boldsymbol{\Sigma}^{-1} (\boldsymbol{\mu}^{(1)} - \boldsymbol{\mu}^{(2)})}$ (Mahalanobis distance between the mean vectors of the two populations). The distance $\Delta$ is the standardized effect size for the multivariate differences between the two populations [3]. The distribution of $s|z$ is given by (adapted from [4]):

$$s|z \sim N_1\left((-1)^z \cdot \frac{\Delta}{2}, 1\right) \quad \text{(S5)}$$

With the distribution in Equation (S4) and given boundary $b$ the expected false positive rate $P(s \geq b|z = 1)$ and false negative rate $P(s < b|z = 2)$ can be computed. Using differential calculus, the boundary $b$ for the statistic $s$ (that minimize the expected costs as in Equation (S2)) can be computed with (proof can be delivered if requested):

$$b = \frac{\log\left(\frac{1-\pi_0}{\pi_0}\right) + \log\left(\frac{c_2^{(1)}}{c_1^{(2)}}\right)}{\Delta} \quad \text{(S6)}$$

As shown below both misclassification rates (and expected misclassification costs in Equation (S3)) are given entirely with the prescribed misclassification cost parameters, prevalence $\pi_0$ and standardized distance between the mean vectors $\Delta$. Consequently, we denote (for a fixed cost structure) the false positive rate as $FP(\pi_0, \Delta)$, false negative rate as $FN(\pi_0, \Delta)$, the specificity as $SP(\pi_0, \Delta)(= 1 - FP(\pi_0, \Delta))$ and the sensitivity as $SE(\pi_0, \Delta)(= 1 - FN(\pi_0, \Delta))$. With Equations (S4) and (S5) the following equations for the expected misclassification rates, specificity and sensitivity can be derived ($\Phi$ is the cumulative distribution function of a univariate standard normal distribution):

$$FP(\pi_0, \Delta) = P(s \geq b|z = 1) = 1 - \Phi\left(\frac{\log\left(\frac{1-\pi_0}{\pi_0}\right) + \log\left(\frac{c_2^{(1)}}{c_1^{(2)}}\right)}{\Delta} + \frac{\Delta}{2}\right) \quad \text{(S7)}$$

$$FN(\pi_0, \Delta) = P(s < b|z = 2) = \Phi\left(\frac{\log\left(\frac{1-\pi_0}{\pi_0}\right) + \log\left(\frac{c_2^{(1)}}{c_1^{(2)}}\right)}{\Delta} - \frac{\Delta}{2}\right)$$

$$SE(\pi_0, \Delta) = 1 - FN(\pi_0, \Delta) = 1 - \Phi\left(\frac{\log\left(\frac{1-\pi_0}{\pi_0}\right) + \log\left(\frac{c_2^{(1)}}{c_1^{(2)}}\right)}{\Delta} - \frac{\Delta}{2}\right)$$

$$SE(\pi_0, \Delta) = 1 - FP(\pi_0, \Delta) = \Phi\left(\frac{\log\left(\frac{1-\pi_0}{\pi_0}\right) + \log\left(\frac{c_2^{(1)}}{c_1^{(2)}}\right)}{\Delta} + \frac{\Delta}{2}\right)$$

### A non-prospective neutral zone classifier

Neutral zone classifiers add a no decision label $NZ$ to the set of possible predicted outcomes and associated costs $c_{NZ}$. We call a neutral zone classifier that assigns output label based on the current evidence solely (without anticipating distribution of future measurements) as non-prospective neutral zone classifiers. For a non-prospective neutral zone classifier denoted by $\delta_{NPNZ}$ based on the measurements $\boldsymbol{y}$ the label $NZ$ is chosen whenever the expected misclassification costs are higher than $c_{NZ}$ (with $c_{NZ} < min(c_2^{(1)}, c_1^{(2)})$). For given measurements $\boldsymbol{y}$ and corresponding posterior probability $\pi$ the no-decision classifiers $\delta_{NPNZ}$ can be derived by comparing expected costs of each classification outcome as follows (assuming $c_{NZ} \neq (1-\pi)c_2^{(1)}$):

$$\delta_{NPNZ} = \begin{cases} 1, & \pi c_1^{(2)} < \min(c_{NZ}, (1-\pi)c_2^{(1)}) \\ NZ, & c_{NZ} < \min(\pi c_1^{(2)}, (1-\pi)c_2^{(1)}) \\ 2, & (1-\pi)c_2^{(1)} \leq \min(EC_{l.k}, \pi c_1^{(2)}) \end{cases}$$

As for the forced-choice classifier in Equation (S2) also the $\delta_{NPNZ}$ can be defined with a test-statistic $T(\boldsymbol{y})$ while two decision boundaries $b_1$ and $b_2$ are needed to define the regions of classification outcome, i.e. [5]:

$$\delta_{NPNZ} = \begin{cases} 1, & T(\boldsymbol{y}) \leq b_1 \\ NZ, & b_1 < T(\boldsymbol{y}) < b_2, \\ 2, & T(\boldsymbol{y}) \geq b_2 \end{cases} \quad \text{(S8)}$$

If the posterior probability $\pi$ is chosen as test statistic, the decision boundaries are (adapted by [1]):

$$b_1 = \frac{c_{NZ}}{c_1^{(2)}}, b_2 = \frac{c_2^{(1)} - c_{NZ}}{c_2^{(1)}} \quad \text{(S9)}$$

In case $b_1 < b_2$, the classifier $\delta_{NPNZ}$ with $\pi$ and corresponding $b_1$ and $b_2$ in equation (S9) minimizes the expected costs, if not, we end up with the forced-choice classifier as minimum cost classifier [1]. Since $c_{NZ} = b_1 c_1^{(2)}$ and $c_{NZ} = (1 - b_2) c_2^{(1)}$, one can see that $\delta_{NPNZ}$ predict the label $NZ$ in case the expected misclassification costs when choosing one of the classes are higher than costs $c_{NZ}$ for not making a decision. Of note, equation (S9) is valid for arbitrary distributions of $\boldsymbol{y}$.

#### Expected cost reduction with optional measurements for prospective neutral zone classifier

For the prospective neutral zone classifier, we differentiate between passed measurements ($\boldsymbol{y}_k$) and optional future measurements ($\boldsymbol{y}_l$). The whole vector of measurements $\boldsymbol{y}$ is associated with measurement costs, whereas we denote the measurement costs of the future measurements $\boldsymbol{y}_l$ with $c^{\mathcal{M}}$ and set in the following the measurement costs of $\boldsymbol{y}_k$ to 0 since it has no influence on the classification outcome of the prospective neutral zone classifier. The expected misclassification costs for a forced choice classifier $\delta_{FC,k}$ based on the already assessed measurements $\boldsymbol{y}_k$ are $EC_k = E(C_k(\boldsymbol{y}_k, \mathrm{z})|\boldsymbol{y}_k) = min(\pi_k c_1^{(2)}, (1 - \pi_k) c_2^{(1)})$ where $\pi_k = P(z = 2|\boldsymbol{y}_k)$ is computed with equation (2) from the main text of the article (by plugging in $\boldsymbol{y}_k$ and the corresponding densities of the two populations) and is called current evidence. Since $\boldsymbol{y}_k$ is given, the expected misclassification rates (and consequently expected costs) based on the whole vector $\boldsymbol{y}$ conditional on $\boldsymbol{y}_k$ depend only on the parameters of the distribution of $z|\boldsymbol{y}_k$ (current evidence $\pi_k$) and the distribution of $\boldsymbol{y}_l|z, \boldsymbol{y}_k$ (given in equation (S1)). The expected total costs (misclassification and measurement costs) of a forced choice classification based on all measurements $\boldsymbol{y}$ conditioning on the already assessed $\boldsymbol{y}_k$ are given by:

$$\begin{aligned} E(C(\boldsymbol{y}, z)|y_k) &= EC_{l.k} \\ &= c^{\mathcal{M}} + FP\big(\pi_k, \boldsymbol{\mu}_{l.k}{}^{(1)}, \boldsymbol{\mu}_{l.k}{}^{(2)}, \boldsymbol{\Sigma}_{l.k}{}^{(1)}, \boldsymbol{\Sigma}_{l.k}{}^{(2)}\big)\,(1 - \pi_k) c_2^{(1)} \\ &\quad + FN\big(\pi_k, \boldsymbol{\mu}_{l.k}{}^{(1)}, \boldsymbol{\mu}_{l.k}{}^{(2)}, \boldsymbol{\Sigma}_{l.k}{}^{(1)}, \boldsymbol{\Sigma}_{l.k}{}^{(2)}\big)\pi_k c_1^{(2)} \end{aligned} \tag{S10}$$

Again, we derived closed form solutions for the expected misclassification rates for the homogeneous case and approximated them with Monte Carlo simulations for the heterogeneous case. Given the current evidence and both misclassification rates the costs $EC_{l.k}$ can be computed. By comparing the expected costs $EC_k$ and $EC_{l.k}$ the prospective neutral zone classifier $\delta_{PNZ,k}$ can be derived. The classifier $\delta_{PNZ,k}$ assigns the label $NZ$ whenever the costs can be reduced with the inclusion of $\boldsymbol{y}_l$. From all possible classification outcomes $\{1; NZ; 2\}$ the prospective sequential neutral zone classifier $\delta_{PNZ,k}$ choose the one with lowest expected costs (see Equation (6) in the main part of the article where only the homogeneous case is discussed but equation also applies to the heterogeneous case). When applying the non-prospective neutral zone classifier with boundaries as in equation (S9) with the current evidence $\pi_k$, the label $NZ$ is predicted independently of the added value of the left-over observations $\boldsymbol{y}_l$. When applying equation (S9) directly for the sequential situation by setting $c^{NZ} = c^{\mathcal{M}}$, it is assumed that when choosing the label $NZ$ no misclassification costs follow afterwards, i.e., that the expected misclassification rates are $FP\big(\pi_k, \boldsymbol{\mu}_{l.k}{}^{(1)}, \boldsymbol{\mu}_{l.k}{}^{(2)}, \boldsymbol{\Sigma}_{l.k}{}^{(1)}, \boldsymbol{\Sigma}_{l.k}{}^{(2)}\big) = FN\big(\pi_k, \boldsymbol{\mu}_{l.k}{}^{(1)}, \boldsymbol{\mu}_{l.k}{}^{(2)}, \boldsymbol{\Sigma}_{l.k}{}^{(1)}, \boldsymbol{\Sigma}_{l.k}{}^{(2)}\big) = 0$. Furthermore, when plugging in the future expected costs $EC_{l.k}$ as neutral costs in equation (S9) (i.e., setting $c^{NZ} = EC_{l.k}$ as random variable rather than a fixed scalar) the fixed points equations discussed later (equations (S16) and (S17)) follow (proof can be delivered if requested).

#### Minimum expected increase in accuracy as threshold for the prospective neutral zone classifier

In this study we set in all analyses $c_1^{(2)} = c_2^{(1)} = 100$ such that the measurement costs $c^{\mathcal{M}}$ of the prospective sequential classifier can be interpreted as the percentage of one misclassification i.e., that $x$ measurements are equally costly as $\frac{x \cdot c^{\mathcal{M}}}{100}$ misclassification. We denote with $A_k$ the expected accuracy for a forced-choice classification based on the passed, completed measurements $\boldsymbol{y}_k$ and with $A_{l.k}$ the (prospective) expected accuracy of a forced-choice classifier based on (unknown) measurements $\boldsymbol{y}_l$ given passed measurements $\boldsymbol{y}_k$. The expected accuracies and consequently increase in accuracy ($dA_{l.k}$) are given by:

$$\begin{aligned} A_k &= max(\pi_k, 1-\pi_k) \\ A_{l.k} &= 1 - \big(FP(\pi_k, \boldsymbol{\mu}_{l.k}{}^{(1)}, \boldsymbol{\mu}_{l.k}{}^{(2)}, \boldsymbol{\Sigma}_{l.k}{}^{(1)}, \boldsymbol{\Sigma}_{l.k}{}^{(2)})\,(1-\pi_k) \\ &\quad + FN(\pi_k, \boldsymbol{\mu}_{l.k}{}^{(1)}, \boldsymbol{\mu}_{l.k}{}^{(2)}, \boldsymbol{\Sigma}_{l.k}{}^{(1)}, \boldsymbol{\Sigma}_{l.k}{}^{(2)})\pi_k\big) \\ dA_{l.k} &= A_{l.k} - A_k \end{aligned} \tag{S11}$$

As described before, the prospective neutral zone classifier chooses the label NZ whenever $EC_{l.k} < EC_k$. For the situation where both misclassification costs are set to 100 these expected costs can be written as $EC_k = 100 \cdot (1 - A_k)$ respectively $EC_{l.k} = c^{\mathcal{M}} + 100 \cdot (1 - A_{l.k})$ such that the condition when the label NZ is chosen by a prospective neutral zone classifier can be re-formulated as (proof with elementary calculus):

$$A_{l.k} - A_k > \frac{c^{\mathcal{M}}}{100} \tag{S12}$$

This means that the optional measurements $\boldsymbol{y}_l$ are only considered for classification in case the accuracy increase expected by their inclusion is higher than $\frac{c^{\mathcal{M}}}{100}$ (e.g., if $c^{\mathcal{M}} = 4$ the measurements $y_l$ are only assessed when accuracy is expected to increase by at least 0.04).

#### Prospective neutral zone classifier for the homogeneous case

For the homogenous case we derived a closed form solutions for the expected misclassification rates using the following test statistic and decision boundary (minimizing the expected costs in equation (S10)):

$$s_{l.k} = \frac{\left(\boldsymbol{y}_l - \boldsymbol{\mu}_{l.k}^{(1,2)}\right)^T \boldsymbol{\Sigma}_{l.k}{}^{-1}\left(\boldsymbol{\mu}_{l.k}^{(1)} - \boldsymbol{\mu}_{l.k}^{(2)}\right)}{\Delta_{l.k}}, s_{l.k}|z, \boldsymbol{y}_k \sim N_{m_l}((-1)^z \cdot \frac{\Delta_{l.k}}{2}, 1)$$

$$b_{l.k} = \frac{\log\left(\frac{1-\pi_k}{\pi_k}\right) + \log\left(\frac{c_2^{(1)}}{c_1^{(2)}}\right)}{\Delta_{l.k}} \tag{S13}$$

where $\boldsymbol{\mu}_{l.k}^{(1,2)} = \frac{\boldsymbol{\mu}_{l.k}^{(1)} - \boldsymbol{\mu}_{l.k}^{(2)}}{2}$ and $\Delta_{l.k} = \left\|\boldsymbol{\mu}_{l.k}{}^{(2)} - \boldsymbol{\mu}_{l.k}{}^{(1)}\right\|_{\boldsymbol{\Sigma}_{l.k}} = \sqrt{(\boldsymbol{\mu}_{l.k}{}^{(1)} - \boldsymbol{\mu}_{l.k}{}^{(2)})^T \boldsymbol{\Sigma}_{l.k}{}^{-1}(\boldsymbol{\mu}_{l.k}{}^{(1)} - \boldsymbol{\mu}_{l.k}{}^{(2)})}$ (similarly as in the Equation (S4) - (S6) for the derivation of the misclassification costs defined in

Equation (S3)). We call $\Delta_{l.k}$ the prospective discriminability which quantifies the added value of the measurements $\boldsymbol{y}_l$ for classification (effect size of differences between sub-populations with given values $y_k$). As before in Equation (S7) but with the current evidence $\pi_k$ as "prevalence" and the distributions and decision boundary in Equation (S13) the expected misclassification rates can be computed. The expected misclassification rates are given (besides the prescribed misclassification costs) as a function of $\pi_k$ and $\Delta_{l.k}$ , i.e.:

$$FP(\pi_k, \Delta_{l.k}) = P(s_{l.k} \geq \ b_{l.k}|y_k, z=1) = 1 - \Phi\left(\frac{log\left(\frac{1-\pi_k}{\pi_k}\right) + log\left(\frac{c_2^{(1)}}{c_1^{(2)}}\right)}{\Delta_{l.k}} + \frac{\Delta_{l.k}}{2}\right)$$

$$FN(\pi_k, \Delta_{l.k}) = P(s_{l.k} < \ b_{l.k}|y_k, z=2) = \Phi\left(\frac{log\left(\frac{1-\pi_k}{\pi_k}\right) + log\left(\frac{c_2^{(1)}}{c_1^{(2)}}\right)}{\Delta_{l.k}} - \frac{\Delta_{l.k}}{2}\right) \quad \text{(S14)}$$

In our implementation we derived the prospective neutral zone classifier by directly comparing the expected costs of every classification outcome and choosing the option with lowest expected costs (see Equation (6) in the main text of the article). Hence, we did not use a test statistic and two decision boundaries to derive regions of different classification outcomes. For the homogenous case, the prospective neutral zone classifier in Equation (6) from the main text of the article can be written as :

$$\delta_{PNZ,k} = \begin{cases} 1, & \pi_k \leq b_1(\Delta_{l.k}) \\ NZ, & b_1(\Delta_{l.k}) < \pi_k < b_2(\Delta_{l.k}) \\ 2, & \pi_k \geq b_2(\Delta_{l.k}) \end{cases}$$

where the equation only holds in case $b_1(\Delta_{l.k}) < b_2(\Delta_{l.k})$ and the boundaries $b_1(\Delta_{l.k})$ and $b_2(\Delta_{l.k})$. For the prospective neutral zone classifier there exists no closed form solutions for boundaries of the current evidence $\pi_k$ (as shown and discussed in [6] for multi-stage classification) We can think of boundaries $b_1(\Delta_{l.k})$ and $b_2(\Delta_{l.k})$ that gives us the decision for a given level of (current) evidence $\pi_k$ as a function of the left-over diagnosis relevant information $\Delta_{l.k}$. In contrast to the non-prospective neutral zone classifier as in Equation (S9) with constant boundaries for $\pi_k$ that only depend on costs parameters, the prospective neutral zone classifier uses the information about the added value of future measurements to determine if a definite decision can be made with the current evidence or not (stay neutral). The lower boundary $b_1(\Delta_{l.k})$ fulfils that $EC_{l.k} = b_2(\Delta_{l.k})c_1^{(2)}$ and is the solution $\tilde{b}_1$ of the equation (derivation can be delivered on request):

$$\tilde{b}_1 = \frac{c^{\mathcal{M}} + c_2^{(1)} FP\left(\tilde{b}_1, \Delta_{l.k}\right)}{c_1^{(2)} SE\left(\tilde{b}_1, \Delta_{l.k}\right) + c_2^{(1)} FP\left(\tilde{b}_1, \Delta_{l.k}\right)} \quad \text{(S15)}$$

while the upper bound $b_2(\varDelta_{l.k})$ fulfils that $EC_{l.k} = (1 - b_2(\Delta_{l.k})\,)c_2^{(1)}$ and is the solution of the equation:

$$\tilde{b}_2 = \frac{c_2^{(1)} SP(\tilde{b}_2, \Delta_{l.k}) - c^{\mathcal{M}}}{c_2^{(1)} SE(\tilde{b}_2, \Delta_{l.k}) + c_1^{(2)} FN(\tilde{b}_2, \Delta_{l.k})} \tag{S16}$$

Similar fix point forms as in the Equation (S15) and Equation (S16) for the boundaries of $\pi_k$ were derived within a multi-stage classification approach[6]. In case $b_1(\Delta_{l.k}) > b_2(\Delta_{l.k})$ the prospective neutral zone classifier acts as a forced choice classifier with the same misclassification costs. For $\Delta_{l.k} \to \infty$ (so that $FP(\pi_k, \Delta_{l.k}) \to 0$ and $FN(\pi_k, \Delta_{l.k}) \to 0$) the boundaries converge to the ones given by the descriptive neutral zone classifier in equation (S9). A visualization of the forced choice as well as the descriptive and prospective neutral zone can be found in **Fig. S1**.

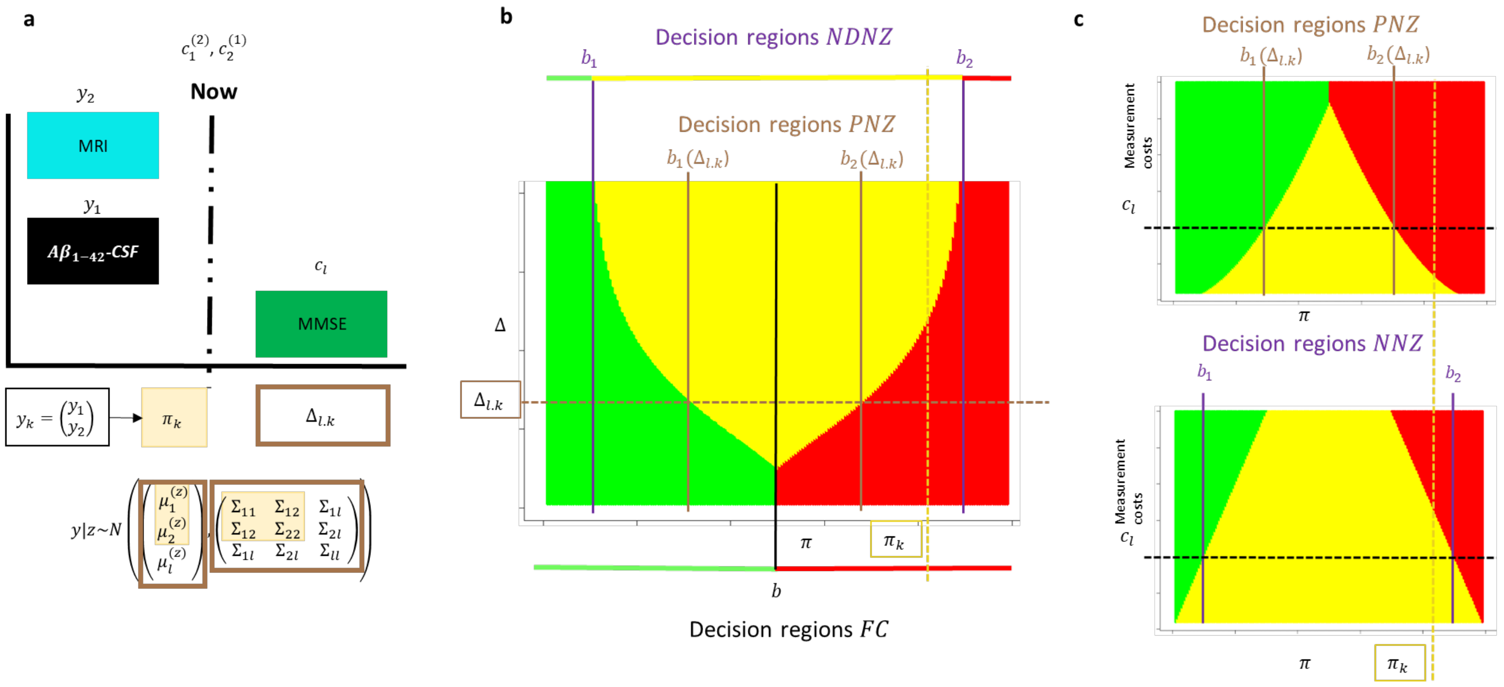

**Fig. S1:** Visualization of the prospective neutral zone classifier **(a)** Situation in which already a MRI ($y_1$) and $A\beta_{1-42}$-CSF ($y_2$) measure were assessed ($\boldsymbol{y}_k$ vector containing both measures) and the classification task consist of either choosing one of the possible classes (MCI-stable or MCI converter) in case there is enough evidence or postpone the decision and stay neutral ($NZ$) to additionally assess the MMSE ($y_l$) and then decide for one of the classes. The classification outcome depends on the misclassification costs $c_1^{(2)}$, $c_2^{(1)}$ and the measurement cost $c_l$ for $y_l$, the current evidence $\pi_k$ and the prospective discriminability $\Delta_{l.k}$ (given by the conditional distributions $y_l|z, \boldsymbol{y}_k$). **(b)** Visualizations of decision regions for constant decision costs. In contrast to the forced-choice (FC, black line) and the non-prospective neutral zone classifier (NPNZ, purple lines) for the prospective neutral zone classifier (PNZ) the amount of leftover diagnosis-relevant information in $\boldsymbol{y}_l$ (quantified by $\Delta_{l.k}$) determines the amount of evidence given by the assessment $\boldsymbol{y}_k$ (quantified by $\pi_k$) that is needed to make a finite decision without assessing $y_l$ ($\pi_k < b_1(\Delta_{l.k})$ or $\pi_k > b_2(\Delta_{l.k})$). For the prospective neutral zone classifier, the prediction outcome for a given value of $\Delta_{l.k}$ is displayed by the coloured areas, the boundaries between the green and yellow area is $b_1(\Delta_{l.k})$ and the one between the yellow and red area is $b_2(\Delta_{l.k})$ **(c)** For fixed misclassification costs $c_1^{(2)}$, $c_2^{(1)}$ and prospective discriminability $\Delta_{l.k}$ the decision boundaries for the PNZ- and DNZ classifiers are displayed as a function of the measurement costs

### Supplementary Methods 3: More information about Linear mixed-effects models

#### Linear mixed effects models for multi-variate, longitudinal data

In this study we used an unbalanced longitudinal data set with a large range of age at study entry and follow-up time period as well as varying time intervals between observations. Under such conditions cross-sectional differences and longitudinal trends may be different and should be estimated simultaneously [7]. A parametrization using baseline age (the age of the subject at the first visit) and the time since baseline (age at each visit minus baseline age) as predictors instead of directly using the age at each visit as model predictors was considered in this study (as in earlier studies to analyze longitudinal data, see [7,8].) We start with the derivation of a model with longitudinal measurements of one response variable in the LMM (univariate case). Let $a_i$ be the baseline age of subject $i$ and $t_{i,j}(\forall i \in \{1,2,\dots,n\}: \forall j \in \{1,2,\dots,m_i\})$ the time since baseline of subject $i$ at the visit $j$. We can model the measurement $y_{i,j}$ using an overall fixed intercept ($\beta_1$), fixed effects for the baseline age ($\beta_2$) and time ($\beta_2$) and random effects per subject for the intercept ($\zeta_{i,1}$ $i \in \{1,2,\dots,n\}$) and for time ($\zeta_{i,2}$ $i \in \{1,2,\dots,n\}$), i.e.:

$$y_{i,j} = \beta_1 + \beta_2 a_i + \beta_3 t_{i,j} + \zeta_{i,1} + \zeta_{i,2} t_{i,j} + \varepsilon_{i,j} = \alpha_i + \lambda_i t_{i,j} + \varepsilon_{i,j} \qquad \text{(S17)}$$

For this model we assume independent and identically distributed residuals $\varepsilon_{i,j} \sim N(0,\rho)$ and a random vector $\boldsymbol{\zeta}_i = \begin{pmatrix}\zeta_{i1}\\ \zeta_{i2}\end{pmatrix} \sim N_2(0,\boldsymbol{\Psi})$ containing random subject-specific deviations $\zeta_{i,1}$ of the intercept and $\zeta_{i,2}$ of the slope over time. The covariance matrix of the random effects $\boldsymbol{\Psi} = \begin{pmatrix}\psi_{11} & \psi_{12}\\ \psi_{12} & \psi_{22}\end{pmatrix}$ consists of the variance of random effects for the intercept ($\psi_{11}$), the variance of the random effects for the slope in time ($\psi_{22}$) and the covariance between the two random effects ($\psi_{12}$). The mean $\boldsymbol{\mu}_i = E(\boldsymbol{y}_i)$ of the vector $\boldsymbol{y}_i$ containing all measurements of subject $i$ is given by the vector of fixed effects $\boldsymbol{\beta}$ (since all random effects and residuals have a mean of 0) and the covariance matrix $Var(\boldsymbol{y}_i)$ by $\boldsymbol{\Psi}$ and $\rho$. We have subject specific random intercepts $\alpha_i = \beta_1 + \beta_2 a_i + \zeta_{i,1}$ consisting of a fixed average (population-level) intercept $\beta_1 + \beta_2 a_i$ for subjects with baseline age $a_i$ and a random deviation $\zeta_{i,1}$ and random slopes in time $\lambda_i = \beta_3 + \zeta_{i,2}$ given by a fixed average slope $\beta_3$ and a random deviation $\zeta_{i,2}$.. The model parameters are the three fixed effects ($\beta_1$, $\beta_2$ and $\beta_2$), the two variances and the covariance of the random effects ($\psi_{11}, \psi_{22}$ and $\psi_{21}$)and one residual variance $\rho$.

Within the LMM framework it is also possible to model repeated measures of multiple response variables simultaneously (multivariate case) [8–10]. Suppose we have $r$ different response variables whereas these responses $h \in \{1;2;\dots;r\}$ are measured $m_{h,i}$ times for subjects $i \in \{1;2;\dots;n\}$ so that for subject $i$ we have overall $m_i = \sum_{h=1}^{r} m_{h,i}$ observations. The number of observations $m_{h,i}$ and the measurement time points can differ between the variables and subjects (19, 21, 25, 26). Let $\boldsymbol{y}_i$ respectively $\boldsymbol{t}_i$ be the vectors containing all measurements respectively measurement time points of subject $i$ (of all responses). To model the measurement $y_{i,j}$ ($j \in \{1;2;\dots;m_i\}$) we additionally to the baseline age $a_i$ and measurement time (since baseline) $t_{i,j}$ include $r$ different dummy variables $v_{h,i,j}$

that are 1 if it is a measurement of the variable $h$ and 0 else as predictor variables. The model looks similar as the one in equation (S17) but for the multivariate case we consider individual fixed and random effects as well as intra-subject variances for every response variable $h \in \{1; 2; \dots; r\}$. To simplify notation, we use in the model equation of the multi-variate case (equation (S18) that follows below) scaled residuals $\epsilon_{i,j}$ instead of the unscaled (raw) residuals $\varepsilon_{i,j}$ (as for the uni-variate case in equation (S17)). The extended model for the multivariate case is given by (adapted from [10–12]):

$$
\begin{aligned}
y_{i,j} &= \sum_{h=1}^{r} v_{h,i,j}\left(\beta_{h,1} + \beta_{h,2}a_i + \beta_{h,3}t_{i,j} + \zeta_{h,i,1} + \zeta_{h,i,2}t_{i,j} + \rho_h\epsilon_{i,j}\right) \\
&= \sum_{h=1}^{r} v_{h,i,j}\left(\alpha_{h,i} + \lambda_{h,i}t_{i,j} + \rho_h\epsilon_{i,j}\right)
\end{aligned}
\qquad \text{(S18)}
$$

with random subject- and response variable specific intercepts $\alpha_{h,i} = \beta_{h,1} + \beta_{h,2}a_{i,1} + \zeta_{h,i,1}$ and slopes in time $\lambda_{h,i} = \beta_{h,3} + \zeta_{h,i,2}$ (given by the fixed and random effects) and scaled residuals $\epsilon_{i,j}$. As can be seen in equation (S18), all considered effects of predictors are interaction effects with the dummy variables $v_{h,i,j}$ ($h \in \{1; 2; \dots; r\}$) such that there are no effects included that effect $y_{i,j}$ in the same way for all considered response variables. To model the means $3r$ parameters i.e., the fixed effects $\beta_{h,1}$, $\beta_{h,2}$ and $\beta_{h,3}$ for all response variables are needed. With $\boldsymbol{\beta}$ the vector containing all these fixed effects is denoted. The variances are modelled by the distribution of the random effects $\zeta_{h,i,1}$ and $\zeta_{h,i,1}$ and the intra-subject variance components $\rho_h (h \in H)$, whereas the covariances between two different responses are given slowly by the distribution of the random effects. The scaled residuals $\epsilon_{i,j}$ were assumed to be independent from each other and the random intercept and slopes and standard normal distributed i.e., $\epsilon_{i,j} \sim N(0,1)$. The distribution of the unscaled residuals $\varepsilon_{i,j}$ varies between response variables and is given as $\varepsilon_{i,j} \sim N(0, \sum_{h=1}^{r} v_{h,i,j}\rho_h)$.With $\boldsymbol{\rho} = \begin{pmatrix}\rho_1\\ \rho_2\\ \vdots\\ \rho_r\end{pmatrix}$ we denote the vector containing all response specific variances $\rho_h (h \in H)$. The distribution of the vector of random effects $\boldsymbol{\zeta}_i = \begin{pmatrix}\zeta_{1,i,1}\\ \zeta_{1,i,2}\\ \zeta_{2,i,1}\\ \zeta_{2,i,2}\\ \vdots\\ \zeta_{r,i,1}\\ \zeta_{r,i,2}\end{pmatrix} \sim N_{2r}(0, \boldsymbol{\Psi})$ (random effects for intercept and time for all variables) is given by a mean of 0, individual variances of every random effect, covariances between random intercepts and slopes of the same or different variables as well as between random slopes of different variables. The parameters of the model in equation (S18) are all fixed effects in $\boldsymbol{\beta}$, all variances and covariances in $\boldsymbol{\Psi}$ and all variances in $\boldsymbol{\rho}$.

Modelling diagnosis specific progressions of multi-variate longitudinal measurements. In equation (S18) we considered a LMM for multivariate, longitudinal data whereas the subjects are assumed to only (systematically) differ in terms of baseline age at study entry ($a_i$, $i \in \{1; 2; \dots; n\}$). In this study we were interested in modelling diagnosis specific progressions disease relevant markers i.e., that the measurements $y_{i,j}$ are coming from populations with differing clinical diagnosis $z \in \{1; 2\}$. Such LMMs can be used to train diagnosis-specific distributions based on labelled measurements denoted by $\boldsymbol{y}_i^{(z_i)}$ ($z_i \in \{1; 2\}$ known diagnosis of subject $i$) that can be used to classify future subjects for which we have access to the measurements but the diagnosis is unknown [9]. To this end we extend the model in equation (S18) by including the diagnosis $z_i$ as a predictor to model the labelled response values $y_{i,j}^{(z_i)}$. The considered model is given by:

$$
\begin{aligned}
y_{i,j}^{(z_i)} &= \sum_{h=1}^{\mathrm{r}} v_{h,i,j} \left( \beta_{h,1}^{(z_i)} + \beta_{h,2}^{(z_i)} a_i + \beta_{h,3}^{(z_i)} t_{i,j} + \zeta_{h,i,1} + \zeta_{h,i,2} t_{i,j} + \rho_h^{(z_i)} \epsilon_{i,j} \right) \\
&= \sum_{h=1}^{\mathrm{r}} v_{h,i,j} \left( \alpha_{h,i}^{(z_i)} + \lambda_{h,i}^{(z_i)} t_{i,j} + \epsilon_{i,j}^{(z_i)} \right)
\end{aligned} \qquad \text{(S19)}
$$

As can be seen in equation (S19) we now have diagnosis specific fixed effects $\beta_{h1}^{(z)}$, $\beta_{h2}^{(z)}$ and $\beta_{h3}^{(z)}$ ($z \in \{1; 2\}$) and assume that they are different for the two populations $z \in \{1; 2\}$. With $\boldsymbol{\beta}^{(z)}$ we denote the vector containing all fixed effects of population $z$ (for all response variables). On the other hand, the random effects $\boldsymbol{\zeta}_i$ are assumed to come from the same distribution, i.e., $\boldsymbol{\zeta}_i \sim N_{2r}(0, \boldsymbol{\Psi})$. The vector of random effects $\boldsymbol{\zeta}_i$ looks the same as before (model in Eq. S14) but now the variances and covariances represent variations only within the same population since the fixed effects account for differences that can be explained by the diagnosis of subjects. Fixed and random effects together give us the random subject specific intercept $\alpha_{h,i}^{(z_i)} = \beta_{h,1}^{(z_i)} + \beta_{h,2}^{(z_i)} a_i + \zeta_{h,i,1}$ and slopes $\lambda_{h,i}^{(z_i)} = \beta_{h,3}^{(z_i)} + \zeta_{h,i,2}$ for all variables $h \in \{1; 2; \dots; r\}$. Now both the diagnosis and baseline age are used to explain between-subject differences in their intercepts. On the other hand, solely the diagnosis is incorporated to explain differences in slopes between subjects. Consequently, the random effects describe differences between average intercepts and slopes for the population the subject belongs to. The random subject-specific intercepts $\alpha_{h,i}^{(z_i)}$ respectively slopes in time $\lambda_{h,i}^{(z_i)}$ are shrinked to the intercept $\beta_{h,1}^{(z_i)} + \beta_{h,2}^{(z_i)} a_i$ respectively slope $\beta_{h,3}^{(z_i)} + \zeta_{h,i,2}$ of the population $z_i \in \{1; 2\}$ the subject $i$ belongs to. As before in equation (S18), $\epsilon_{i,j}$ are the scaled residuals with standard normal distribution but for the model in Eq. S15 the response variable specific intra-subject variances are allowed to differ for the two populations (i.e., differing uncertainty depending on the diagnosis). We implemented two different models assuming that populations with different clinical diagnoses $z$ either (a) only differ in the means (homogenous model) or (b) differ in the means as well as variances (heterogeneous model). For homogenous model, the intra-subject variance components are constrained to be equal between the populations i.e. $\rho_h = \rho_h^{(1)} = \rho_h^{(2)}$ such that unscaled residual are distributed as $\varepsilon_{i,j} \sim N(0, \sum_{h=1}^{r} v_{h,i,j} \rho_h)$ (the same model as the one in equation (8) in the main text). For the heterogeneous model it is assumed that $\rho_h^{(1)} \neq \rho_h^{(2)}$ such that the distribution of the unscaled residuals $\varepsilon_{i,j}$ (respectively their variance) depend on the diagnosis $z_i$ of the subject and is given by $\varepsilon_{i,j} \sim N(0, \sum_{h=1}^{r} v_{h,i,j} \rho_h^{(z_i)})$. All results in the main text of the article are based on homogenous models, whereas the Supplementary Material also cover results of models considering heterogeneous populations (see below).

### Supplementary Methods 4: A multi-variate quadratic, longitudinal discriminant models for non-sequential and sequential classification

#### Generalization to quadratic discriminant model

In the main text of this article, we only considered linear discriminant models (equation (6) given by the LMM in equation (7) from the main text of the article) but in this study, we also embedded mixed-effects modelling within a quadratic discriminant analysis approach (with the more general LMM as in Equation (S19)). In the heterogeneous case we assumed for a subject $i$ with unknown label $z_i$ that

$$z_i \sim Bernoulli(\hat{\pi}_{i,0})$$
$$\boldsymbol{y}_i | z \sim N_{m_i}(\widehat{\boldsymbol{\mu}}_i^{(z)}, \widehat{\boldsymbol{\Sigma}}_i^{(z)}) \tag{S20}$$

Whereas contrast to the linear discriminant approach described in the main article, we have now different covariance matrices $\widehat{\boldsymbol{\Sigma}}_i^{(1)} \neq \widehat{\boldsymbol{\Sigma}}_i^{(2)}$ $(i \in \{1; 2; \dots; n\})$. The estimated densities of the distributions $N_{m_i}(\widehat{\boldsymbol{\mu}}_i^{(z)}, \widehat{\boldsymbol{\Sigma}}_i^{(z)})$ are denoted by $\hat{\phi}_i^{(z)}$ $(z \in \{1; 2\})$. Besides discriminant models with constant prevalence for all participants i.e., assuming $\pi_{i,0} = \pi_0 (\forall i \in \{1, \dots, n\})$ we also considered the case of subject-specific prevalence computed as a function of subject characteristics (that are constant for all observations of a subject). In case a constant prevalence $\pi_0$ is assumed for all subjects, $\pi_0$ was estimated with the relative frequency of participants with diagnosis 2 in the training data. To model subject-specific prevalences, we implemented a logistic regression (as in a previous study[8] that implemented mixed-effects model based discriminant models) assuming that the true prevalence $\pi_{i,0}$ is

$$\pi_{i,0} = P(z_i = 2) = \frac{1}{1 + e^{-(\gamma_0 + \gamma_1 a_i)}} \tag{S21}$$

Our implementation for discriminant models in Equation (S20) had the model parameters $\boldsymbol{\theta} = [\pi_0; \boldsymbol{\beta}^{(1)}; \boldsymbol{\beta}^{(2)}; \boldsymbol{\Psi}; \boldsymbol{\rho}^{(1)}, \boldsymbol{\rho}^{(2)}]$ ($1 + 9r + r^2$ parameters for the heterogeneous and $1 + 8r + r^2$ for the homogenous model with $\boldsymbol{\rho}^{(1)} = \boldsymbol{\rho}^{(2)}$) respectively $[\gamma_0; \gamma_1; \boldsymbol{\beta}^{(1)}; \boldsymbol{\beta}^{(2)}; \boldsymbol{\Psi}; \boldsymbol{\rho}^{(1)}, \boldsymbol{\rho}^{(2)}]$ ($2 + 9r + r^2$ parameters for the heterogeneous and $2 + 8r + r^2$ for the homogenous model). . The parameters $\gamma_0$ and $\gamma_1$ (see equation (S21)) are estimated by fitting (standard) logistic regression given in equation (S21) (one observation per subject, assuming independence between observations) and $\boldsymbol{\beta}^{(1)}$, $\boldsymbol{\beta}^{(2)}$, $\boldsymbol{\Psi}$, $\boldsymbol{\rho}^{(1)}$ and $\boldsymbol{\rho}^{(2)}$ by fitting the LMM given in equation (S19).

All parameters in $\boldsymbol{\theta}$ were estimated with training data with labelled observations using a 20-fold cross validation framework. We randomly split the sample into 20 subsamples (folds) and used for the classification of a subject coming from a fold $f$ the parameters estimated with the combination of all remaining folds. Using the training data consisting of 19 folds, we estimated all parameters (prevalence and LMM parameters) denoted by $\widehat{\boldsymbol{\theta}}_{-f}$. Given the parameter estimates $\widehat{\boldsymbol{\theta}}_{-f}$ for a subject $i$ from fold $f$ we computed the estimators $\widehat{\boldsymbol{\mu}}_i^{(z)}$ and $\widehat{\boldsymbol{\Sigma}}_i^{(z)}$ from all $m_i$ observations of a subject $i$ ($i \in \{1; 2; \dots; n\}$). With the estimated mean vectors and covariance matrices we computed all previously derived quantities (posterior probabilities, expected miss-classification rates, expected costs and classifiers) by plugging in the estimates $\hat{\pi}_{i,0}$, $\widehat{\boldsymbol{\mu}}_i^{(z)}$ and $\widehat{\boldsymbol{\Sigma}}_i^{(z)}$ (or a subset of components thereof) of subject $i$ for the true population values in the formula.

Within our library POSEIDON (https://git.upd.unibe.ch/openscience/POSEIDON.git) the (linear and quadratic) discriminant model based on mixed-effects model estimation can be fitted with the function *daLmeMulti*. Based on a trained model and a data set of one subject containing all needed predictor values (i.e., in our study: dummy variables for marker identification, age at baseline of the subject and the time since baseline for every visit, see Equations (S19) and (S21)) the mean vectors $\widehat{\boldsymbol{\mu}}_i^{(z)}$ and covariance matrices $\widehat{\boldsymbol{\Sigma}}_i^{(z)}$ of the marker values can be predicted with the function *predDistParamLme.* More information about the software can be found in the main article or especially in the vignette "*trainingInferenceClassificationLmm*" that is included in the POSEIDON library.

### A sequential classification framework

In this study we derived sequential classifiers that predicts for a subject $i$ at the step $k$ ($1 \leq k < m_i$) one of the possible diagnoses or stay in the neutral zone (forced-choice classification in case $k = m_i$). Let $M_{i,k} \subset \{1; 2; \dots; m_i\}$ be the set of all $k$ indices of the measurements that are already assessed, $\boldsymbol{y}_{i,k} = \left(y_{i,j}\right)_{j \in M_{i,k}}$ the vector containing all passed observations and $\hat{\pi}_{i,k}$ the estimated current evidence for subject $i$ at step $k$ (plugging in values $\boldsymbol{y}_{i,k}$ and estimated prevalence $\hat{\pi}_{i,0}$in equation (2) in main text of this article). We define $\widetilde{M}_{i,k}$ as the set of indices of left-over observations i.e., of observations with indices not contained in $M_{i,k}$ and assessed not before $t_{i,k}^{\max}$ with $t_{i,k}^{\max} = \max_{j \in M_{i,k}} \left(t_{i,j}\right)$. Using the estimated distributions of $y_{i,l}$ (with $l \in \widetilde{M}_{i,k}$) conditional on $z$ and $\boldsymbol{y}_{i,k}$ (can be computed by plugging in the estimated mean and covariances in equation (S1)) we estimated the expected costs $\widehat{EC}_{i,l.k}$ by first estimating the expected false positive and false negative rates and then plug them in equation (S14). Of note, for the first application we only consider two measurements, for $k = 1$ we apply a prospective neutral zone classifier as in equation (5) from the main text and for $k = 2$ (second stage) we already choose one of the possible classes. Hence, for this application we have for $k = 1$ one passed measurement $y_{i,j_1} \in \mathbb{R}$ as well as one leftover measurement $y_{i,j_2} \in \mathbb{R}$. In the following we first describe the computation of estimators of the expected false positive and false negative rates for both the homogeneous and heterogeneous case (needed for the first and second application of this study) and then describe our novel algorithm to sequentially select longitudinal sequence of multiple markers (second application of this study).

To get the estimated misclassification rates for a classification with a left-over measurement $y_{i,l}$ ($l \in \widetilde{M}_{i,k}$) given the values of the already assess measurements $\boldsymbol{y}_{i,k}$ we computed in the homogeneous case (linear discriminant model) the estimated prospective discriminability $\widehat{\Delta}_{i,l.k} =$

$\sqrt{\left(\widehat{\boldsymbol{\mu}}_{i,l.k}^{(1)}-\widehat{\boldsymbol{\mu}}_{i,l.k}^{(2)}\right)^T \boldsymbol{\Sigma}_{i,l.k}{}^{-1}\left(\widehat{\boldsymbol{\mu}}_{i,l.k}^{(1)}-\widehat{\boldsymbol{\mu}}_{i,l.k}^{(2)}\right)}$ and plugged it in together with the estimated current evidence $\hat{\pi}_{i,k}$ in Equation (S14) to get estimates for the expected misclassification rates (with which we can compute the expected costs as in equation (S10)). The estimation of the expected misclassification rates of a measurement $y_{i,l}$ ($l \in \widetilde{M}_{i,k}$) given the already assessed measurements $\boldsymbol{y}_{i,k}$ in the heterogenous case (quadratic discriminant model) involved Monte Carlo simulations. To approximate the misclassification rates we considered the current evidence $\hat{\pi}_{i,k}$ and the distributions $N_1\left(\hat{\mu}_{i,l.k}^{(z)}, \hat{\Sigma}_{i,l.k}^{(z)}\right)$ ($\hat{\mu}_{i,l.k}^{(z)}$ estimator for $E\left(y_{i,l} \middle| \boldsymbol{y}_{i,k}, z\right)$ and $\hat{\Sigma}_{i,l.k}^{(z)}$ for $Var\left(y_{i,l} \middle| \boldsymbol{y}_{i,k}, z\right)$) with corresponding density function estimate $\hat{\phi}_{i,l.k}{}^{(z)}$. We denote with $y_{s,l.k}^{(z)}$ the value of a simulation $s$ from the distribution $N_1\left(\hat{\mu}_{i,l.k}^{(z)}, \hat{\Sigma}_{i,l.k}^{(z)}\right)$ ($z \in \{1; 2\}$). As test-statistic for classification we computed for every simulation $\hat{\pi}_{s,l.k}^{(z)}$ by plugging in $\hat{\phi}_{i,l.k}{}^{(1)}\left(y_{s,l.k}^{(z)}\right)$ respectively $\hat{\phi}_{i,l.k}{}^{(2)}\left(y_{s,l.k}^{(z)}\right)$ and the estimated current evidence $\hat{\pi}_{i,k}$ (instead of the prevalence $\pi_0$) in equation (2) from the main part of this article. Specifically, to compute the estimated misclassification rates we implemented the following procedure: (a) For $z \in \{1; 2\}$ we simulated 1000 values $y_{s,l.k}^{(z)}$ ($s \in \{1; 2; \dots; 1000\}$) from $N_1\left(\hat{\mu}_{i,l.k}^{(z)}, \hat{\Sigma}_{i,l.k}^{(z)}\right)$ and (b) computed for every simulation the test-statistic $\hat{\pi}_{s,l.k}^{(z)}$, (c) with the overall 2000 simulations we estimated the expected false positive respectively negative rate as

$$
\begin{aligned}
FP\left(\pi_k, \widehat{\boldsymbol{\mu}}_{i,l.k}^{(1)}, \widehat{\boldsymbol{\mu}}_{i,l.k}^{(2)}, \widehat{\boldsymbol{\Sigma}}_{i,l.k}^{(1)}, \widehat{\boldsymbol{\Sigma}}_{i,l.k}^{(2)}\right) &= \frac{\#\left\{s\colon \hat{\pi}_{s,l.k}^{(1)} \geq \dfrac{c_2^{(1)}}{c_2^{(1)}+c_1^{(2)}}\right\}}{1000} \\
FN\left(\pi_k, \widehat{\boldsymbol{\mu}}_{i,l.k}^{(1)}, \widehat{\boldsymbol{\mu}}_{i,l.k}^{(2)}, \widehat{\boldsymbol{\Sigma}}_{i,l.k}^{(1)}, \widehat{\boldsymbol{\Sigma}}_{i,l.k}^{(2)}\right) &= \frac{\#\left\{s\colon \hat{\pi}_{s,l.k}^{(2)} < \dfrac{c_2^{(1)}}{c_2^{(1)}+c_1^{(2)}}\right\}}{1000}
\end{aligned}
\qquad \text{(S22)}
$$

For sequential classification with longitudinal sequences we computed for every left-over observation $l \in \widetilde{M}_{i,k}$ of subject $i$ at the step $k$ the measurements costs $c_{i,l}^{\mathcal{M}} = c_t\left(t_{i,l} - t_{i,k}^{\max}\right) + \sum_{h\in H} c_h v_{h,i,l}$ and the estimators for the misclassification rates (as described above). By plugging in the estimated current evidence $\hat{\pi}_{i,k}$, false positive and false negative rates and measurement costs $c_{i,l}^{\mathcal{M}}$ in equation (S10) we got the estimated expected costs by the inclusion of observation $l$ given the already assessed observations in $M_{ik}$ denoted by $\widehat{EC}_{i,l.k}(l \in \widetilde{M}_{i,k})$. Given the expected costs $\widehat{EC}_{i,l.k}$ ($l \in \widetilde{M}_{i,k}$) of all leftover observations $l \in \widetilde{M}_{i,k}$ we formulated decision and selection rules to derive the sequential algorithm. We constructed the sequential classifier $\hat{\delta}_{seq,i,k}$ that assigns at a step $k$ the outcome labels as follows (assuming $\left(1-\hat{\pi}_{i,k}\right)c_2^{(1)} \neq \min\limits_{l\in\widetilde{M}_{i,k}}\left(\widehat{EC}_{i,l.k}\right)$):

$$\hat{\delta}_{seq,i,k} = \begin{cases} 1, & \hat{\pi}_{i,k} c_1^{(2)} \le \min\left( \min_{l \in \widetilde{M}_{i,k}} \left( \widehat{EC}_{i,l.k} \right), (1-\hat{\pi}_{i,k}) c_2^{(1)} \right) \\ NZ, & min_{l \in \widetilde{M}_{i,k}} \left( \widehat{EC}_{i,l.k} \right) < \min\left( \hat{\pi}_{i,k} c_1^{(2)}, (1-\hat{\pi}_{i,k}) c_2^{(1)} \right) \\ 2, & (1-\hat{\pi}_{i,k}) c_2^{(1)} < \min\left( \min_{l \in \widetilde{M}_{i,k}} \left( \widehat{EC}_{i,l.k} \right), \hat{\pi}_{i,k} c_1^{(2)} \right) \end{cases} \quad \text{(S23)}$$

Note, for the sequential classifier we can think of applying the prospective neutral zone classifier in equation (5) from the main text of the article for every left-over observation separately and assigning the label $NZ$ if at least for one observation the prospective neutral zone classifier reveals the label $NZ$ as outcome. The sequential classification algorithms stops if $\hat{\delta}_{seq,i,k} \in \{1; 2\}$, whereas in case $\hat{\delta}_{seq,i,k} = NZ$ a selection rule is applied to choose which (single) observation $l^*$ is included next for the prediction. Let $\widehat{EC}_{i,k}$ be the expected cost for a classification with the current evidence $\hat{\pi}_{i,k}$ such that $\widehat{EC}_{i,k} - \widehat{EC}_{i,l.k}$ is the expected cost reduction by including observation $l$. We used two different selection rules i.e., the greedy rule where the earliest observation with expected cost reduction or the exhaustive rule where the observations with highest expected cost reduction is chosen as the next observation $l^*$. In case of multiple observations with expected cost reductions at the earliest possible time, the greedy rule chose the observations with highest expected cost reduction. Within the exhaustive rule, the earlier observation is chosen if there are multiple observations with expected cost reduction that are equal to the highest possible reduction. Afterwards the sequential algorithm continues by setting $M_{i,k+1} = M_{i,k} \cup \{l^*\}$ until it stops at a step $K_i \le m_i$ when no observation with expected cost reduction or no left-over observation are available such that $\hat{\delta}_{SNZ,i,K_i} \in \{1; 2\}$.

In our library POSEIDON (https://git.upd.unibe.ch/openscience/POSEIDON.git) algorithms for prospective sequential diagnosis with neutral zones are implemented. The library allows to apply two-stage classifiers (function "*twoStageClass*") as well as sequential classifiers to include longitudinal sequences for prediction (function "*seqLongClass*") for given distributions estimates (given by the function *predDistParamLme* of POSEIDON). More information about the statistical implementation of sequential classifiers can be found in the main article of this study or especially in the vignette "*trainingInferenceClassificationLmm*" (included in the library POSEIDON.).

### Supplementary Methods 5: Time-to-event analyses to analyse conversion times

In this study we also performed time to event analyses using the time until clinical manifestation of AD (conversion time) considering MCI-stables as right censored data. We compared positive and negative predicted cases that were labelled either as easy or initially uncertain with a prospective sequential (two-stage) classifier based on a cross-sectional MRI measurement (see main manuscript of this study for more information). We estimated both survival curves (assessing the fraction of not converted participant as a function of time) and hazard ratios for the conversion times. All time-to-event analyses were performed with the R library *survival* [13] using the functions *survfit*, *survdiff* and *coxph.*

The survival curves were estimated using the Kaplan-Meier technique and Independent groups were compared with log-rank (Mantel-Haenzel) significance tests considering a significance level of 0.05. Moreover, hazard ratios were estimated with Cox proportional hazards regression models). Models with the factors subsample (easy or uncertain cases with MRI) and classification (positive or negative predicted cases) were fitted considering effects for both factors and their interaction. We implemented two models with this structure and for both models the classification of a positive or negative label were based on MRI only for all easy case but for one model the classifications of uncertain cases were performed with MRI only and for the other model with both MRI and $A\beta_{1-42}$ measurements. We used the Wald's method to implement significance tests with a significance level of 0.05 and 95% confidence intervals for hazard ratios. The 95% Wald confidence intervals were computed as described in [14]. From model output of the function *coxph* we directly had access to logarithmic hazard ratios ($\hat{\eta}'s$), their standard error ($\widehat{se}(\hat{\eta})'s$) and p-values of the Wald test statistic. We computed the 95% confidence intervals for the hazard ratios as:

$$\left[e^{\hat{\gamma}-1.96\cdot\widehat{se}(\hat{\gamma})}, e^{\hat{\gamma}+1.96\cdot\widehat{se}(\hat{\gamma})}\right] \quad (S24)$$

### Supplementary Material 1: Sample selection

In this study we considered subjects with label mild cognitive impairment (MCI), i.e., patients at risk to develop Alzheimer's disease (AD), and we were interested in separating the ones that stay stable with the MCI label from the one that convert to manifest AD. In the supplementary **Fig. S2** you can find more information about the process of the data set selection. All measurements before the first MCI label were excluded from the analyses. To have a sample of MCI subjects that can be considered as stable we only included subjects that after the first time labelled as MCI stay with this label until the end of the observations period. From these subjects we also excluded the ones that were observed for less than 2.75 years follow-up. On the other hand, we built a sample with subjects that were diagnosed as MCI at a visit and then with the label AD at a later visit, whereas after the first MCI label they have to consistently be labelled as MCI until the first AD diagnosis and then stay with the AD diagnosis until the end of the observation period. Moreover, we only included the subjects that convert to AD within 3.25 years in our sample of MCI converters.

For the training of the models, we only considered subjects with at least eight measurements of either the MMSE, RAVLT, SPARE-AD score or $A\beta_{1-42}$ (number of observations from all markers together was relevant). Overall, a sample of 612 subjects was used for model training.. Moreover, the decision processes of either the first or second application were compared using the same sample (see main text for the description of the used decision processes). Overall 410 subjects were included for the evaluation of the first application according to the criteria: at least one MRI and $A\beta_{1-43}$ within three months from the first observations, while at least one $A\beta_{1-43}$ was not observed before the first MRI measure. For the second application most constraints for inclusion are given by the so called multivariate cross-sectional strategy, since subjects need to have at least one observation of all four markers, whereas all markers have to be assessed within three months after one of these markers was observed for the first time for a subject. Consequently, we used a sample of 403 subjects for the evaluation of the second application.  There was one subject that was included in the evaluation but not training set. For this subject we randomly chose one out of the 20 models.

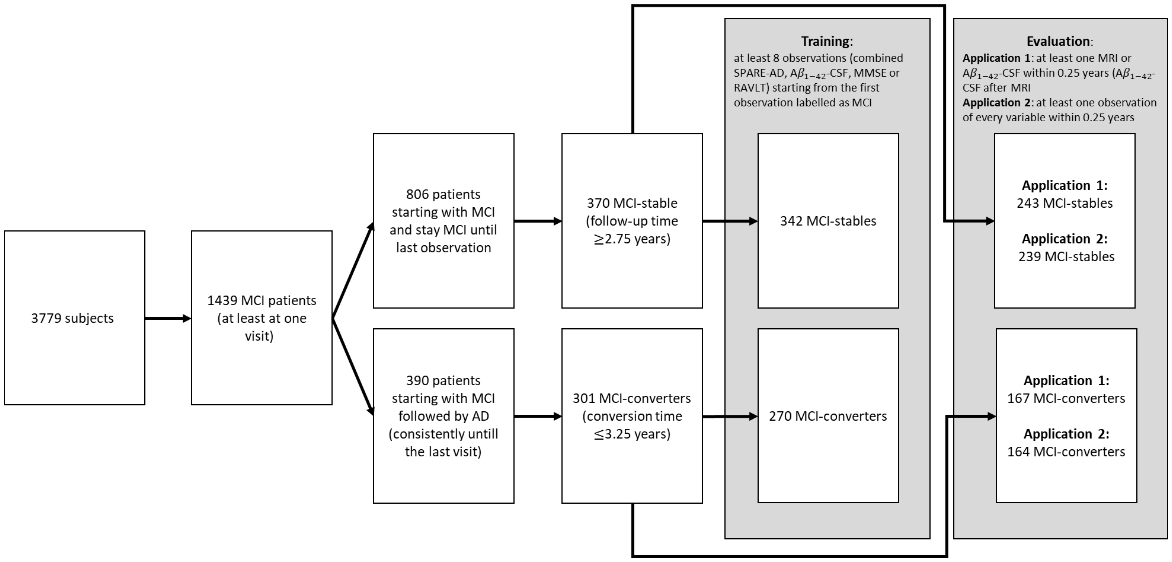

**Fig. S2:** Flow-chart covering the selection of the samples used either for training of model parameters or the evaluation of out-of-sample predictions of different decision processes. We used sequential and fixed (non-sequential) decision strategies to choose the which observations are used for prediction covering two different applications (see main article for more information). We compared the decision processes of either the first or the second application always using the same sample for evaluation. We only considered subjects for evaluation that were also included in training, whereas the training of the models also covered the data of subject that were not included for evaluation (203 in the first and 210 subject from training in the second application were excluded for evaluation).

### Supplementary Results 1: Additional results for two stage classifier

In this section all results covering the first application (two-stage classification) that were not shown in the main text are included, i.e., examination of falsely classified MCI-stables and time-to event analyses (with the model structure used for all results in the main text),

### Examine the raw data of falsely classified MCI-stables

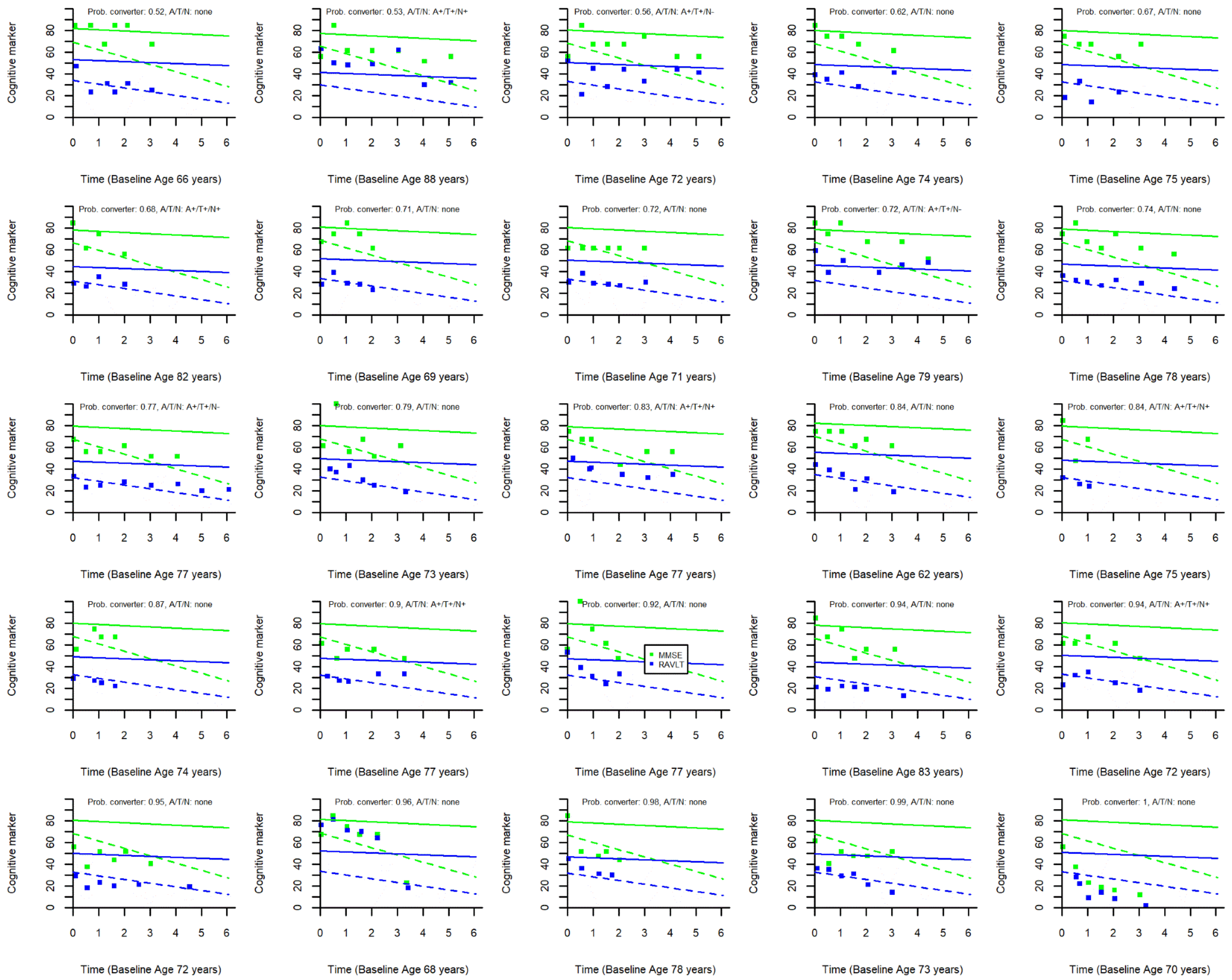

**Fig. S3:** Baseline A/T/N and longitudinal cognitive profiles of false positive cases. The cases got first falsely classified as MCI-converter based on cross-sectional MRI and Amyloid measurement and then also with the cross-sectional MRI and Amyloid and all longitudinal cognitive (MMSE and RAVLT) measurements (see main text of this article for more information). The cases are order according to the predicted probability (out-of-sample via 20-fold cross-validation) of belonging to the population of MCI-converters based on the trained discriminant model used in the main text of the article. Straight lines are the expected average progressions of MCI-stables by the discriminant model (one-fold chosen for the computation), while dashed line are the expected average progression of MCI-converters.

### Time to event analyses with conversion time

Furthermore, we analysed the time from study entry to onset of manifest AD (right censored) using survival curves (Kaplan-Meier estimates) and hazard ratios (estimated with Cox regressions, see Supplementary Methods 5). As shown in **Fig. 5B-C** in the main text of this article, predictions based on

MRI only led to more distinct survival curves respectively higher differences in the hazard rates when fitted on easy cases than when fitted on uncertain cases given by: (a) a steeper survival curve respectively higher hazard rate for easy cases predicted as MCI-converters (non-significant difference p=0.066 respectively p=0.066) but (b) a flatter curve respectively lower hazard rate for easy cases predicted as MCI-stables (significance difference: p<0.001 respectively p<0.001). When additionally, also the A$\beta_{1\text{-}42\text{-}}$ CSF measure is included for uncertain cases the survival curves respectively hazard rates of the ones predicted as MCI-converter and the ones predicted as MCI-stables become more like the ones predicted for easy cases based on MRI only (still significant differences for the ones predicted as MCI-stables: p<.001 respectively p<.001; non-significant differences for the ones predicted as MCI-converters: p=.499 respectively p=.504). For both easy and uncertain cases the survival curves were steeper when they were classified as MCI-converters (see **Fig. 5B**). There was no significant difference between the ssurvival curves fitted on uncertain cases classified as MCI-converters with MRI and the ones fitted on uncertain cases classified as MCI-stables with MRI (p=.136), whereas the difference between the survival curves was highly significant for classifications of easy cases with MRI or classifications of uncertain cases with MRI and A$\beta_{1\text{-}42}$ –CSF. As visualized in **Fig. 5D** (with 95% confidence intervals), the ratios of hazard rates of cases predicted as MCI-converters divided by the one of cases predicted as MCI-stable were not significantly different form one for classifications of uncertain cases with MRI (p=.0.148) but significantly above one for classifications of easy cases with MRI or uncertain cases with MRI and A$\beta_{1\text{-}42\text{-}}$ CSF (p<.001 for both). The hazard ratio in the group of easy cases classified with MRI only was 8.5 times higher than the hazard ratio in the group of uncertain cases classified with MRI only (p<.001) and 3.5 times higher than the hazard ratio in the group of uncertain cases classified with MRI and A$\beta_{1\text{-}42\text{-}}$ CSF (p<.001) (see **Fig. 5D**).

### Supplementary Results 2: Sequential selection of longitudinal sequences

In this section we present on results for non-sequential and sequential classification strategies for a wide range of cost parameters either based on the same model structure considered in the main text or other model structures. All results are included in the Excel file "tabS1.xlsx", here in the Supplementary text only the caption is included.

**Tab. S1**: Multi-objective evaluation of non-sequential and sequential decision strategies for varying cost parameters (see main text for more information). The left part of the table specifies the marker specific costs of acquisition and cost per year of waiting. The first cost parameters are the default costs and for the following parameters deviations from the default costs are highlighted in grey. All objective metrics are based on out-of-sample predictions via 20-fold cross validation. We considered the following metrics (in braces are the column names in the table).

*Summary metric*: mean total costs (tot costs)

*Performance metrics*: Log-loss score (log-loss), accuracy (Acc), Specificity (Spec), Sensitivity (Sens)

*Resource metrics* (marker independent): mean measurement costs (meas cost), mean number of observations (nr. obs), fraction of participants with at least two observations (fraction mind. 2 obs), mean follow-up time (time)

*Marker-specific metrics* ($x$ Name of marker, $x \in \{\mathrm{MMSE}, \mathrm{RAVLT}, \mathrm{SPARE-AD}, \mathrm{Amyloid}\}$): mean number of measurements of marker x (nr x), fraction of participants with at least one measurement of marker x (fraction min. 1 x), mean time of first measurement of marker x (min time x), mean time of the last measurement of marker x (max time x).

The table consists of different sheet. Every sheet contains the results based on another model structure.

(a): (sheet name "a_LDA_predPerv") linear discriminant model with marker model as in Equation (7) in the main text and prevalence model as in the Supplementary Equation (S21)

(b): (sheet name "b_LDA_relFreq") linear discriminant model with marker model as in Equation (7) in the main text and the relative frequency as estimate for the prevalence (constant for all subjects)

(c): (sheet name "c_QDA_predPerv") quadratic discriminant model based on marker model as in the Supplementary Equation (S19) and prevalence model as in the Supplementary Equation (S21)
